# A within-host-informed event-time framework linking epidemiological delay distributions

**DOI:** 10.64898/2026.09.21.26363521

**Authors:** Nyall Jamieson, Sang Woo Park, Kylie Ainslie, Jingsi Xu, Sebastian Funk, Thomas Ward, Christopher E. Overton, Lauren Ancel Meyers, Sam Abbott

**Affiliations:** Department of Integrative Biology, The University of Texas at Austin, Austin, Texas, United States of America; School of Biological Sciences, Seoul National University, Seoul, Korea; Institute for Data Innovation in Science, Seoul National University, Seoul, Korea; Department of Infectious Diseases, The University of Melbourne at the Peter Doherty Institute for Infection and Immunity, Melbourne, Victoria, Australia; School of Public Health, University of Hong Kong, Hong Kong SAR; Department of Mathematics, The University of Manchester, Manchester, United Kingdom; Centre for Mathematical modeling of Infectious Diseases, London School of Hygiene and Tropical Medicine, London, United Kingdom; Data Analytics and Surveillance, UK Health Security Agency, London, United Kingdom; Department of Mathematical Sciences, University of Liverpool, Liverpool, United Kingdom; Department of Statistics and Data Sciences, The University of Texas at Austin, Austin, Texas, United States of America

## Abstract

Understanding when and how pathogens spread from person to person is fundamental to characterizing transmission, interpreting surveillance data, and designing effective control measures. Yet key epidemiological delays—including the latent period, incubation period, generation interval, and serial interval—are difficult to estimate because key events such as infection, infectiousness, symptom onset, and transmission are rarely observed directly. Current approaches typically estimate these delays using flexible statistical distributions rather than explicitly linking them through the underlying processes that generate them, making it difficult to interpret the underlying biology and to ensure consistency across related delay estimates. Here, we develop a stochastic event-time framework built around a dynamic within-host pathogen-growth model that treats epidemiological delays as coupled outcomes of a shared biological process. By representing these key events as linked stochastic events, the framework produces internally consistent delay distributions while inferring parameters with mechanistic interpretations conditional on the assumed within-host model that govern pathogen growth, clearance, infectiousness, and symptom onset. The framework also accommodates censored and partially observed infection and symptom-onset times through a unified observation model. Across simulation studies, the framework accurately recovered generation-interval characteristics across sample sizes and disease time scales. In applications to SARS-CoV-2 and mpox transmission-pair data, the framework showed improved predictive fit relative to conventional lognormal delay models in several analyses, although the magnitude of improvement varied across datasets and observation models; it estimated shorter generation intervals for Omicron than Delta and revealed alternative within-host parameters compatible with the mpox transmission data. This mechanistic framework provides a biologically interpretable alternative to independently fitted delay distributions, yields internally consistent estimates of epidemiological delays, and establishes a foundation for integrating biological and epidemiological data to better understand, predict, and control pathogen transmission.

**Author summary:** For many infections that spread directly from person to person, the timing of infection, infectiousness, symptom onset, and passing the infection to someone else is central to understanding how disease spreads. These delays are often estimated separately using statistical distributions, even though they are biologically connected. For example, the amount of pathogen in the body can influence when someone becomes infectious, when symptoms appear, and when transmission is most likely. We developed a modeling framework that links these timings through a shared pathogen growth process. In the model, becoming infectious, developing symptoms, and transmitting infection occur as chance events whose rates change with pathogen load. We tested the framework using simulations and data from recent SARS-CoV-2 and mpox epidemics, showing that it outperformed conventional statistical approaches and can incorporate pathogen-load data into delay estimation, while providing new estimates of the generation intervals of recent COVID-19 variants and mpox. More broadly, the framework provides a way to combine epidemiological data with biological information, helping researchers understand how changes in within-host (WH) pathogen dynamics influence transmission and improving our ability to predict and control infectious disease outbreaks.

## 1 Introduction

The spread of infectious diseases is shaped by the timing of infection, infectiousness, symptom onset, and onward transmission. Four key epidemiological delays describe these processes: the latent period (infection to infectiousness), the incubation period (infection to symptom onset), the generation interval (infection in a primary case to infection in a secondary case), and the serial interval (symptom onset of a primary case to symptom onset of a secondary case) [1, 2]. These delays are difficult to estimate because the underlying infection and symptom-onset times are often only partially observed or known within intervals [3]. Accurately estimating these delays is important because they determine how transmission unfolds over time and therefore influence estimates of epidemic growth, the timing of control measures, and assessments of intervention effectiveness [4–6]. Although these delays describe different phases of infection and transmission, they are linked by shared within-host biological processes and therefore should not be expected to vary independently [5, 6].

In practice, however, epidemiological delays are commonly modeled as separate marginal distributions, often using flexible gamma, Weibull, or lognormal families [2, 7–9]. Although these distributions can provide useful empirical descriptions and, in some cases, heuristic time-to-event interpretations [10], existing approaches often specify epidemiological delay distributions parametrically, either separately or within joint models, without explicitly linking them to a common set of underlying event times. As a result, estimates of related delays need not be mutually consistent, and similar fitted marginal distributions can imply different underlying timings of infectiousness onset, symptom onset, or transmission.

Mechanistic models provide an alternative by explaining observed epidemiological delays in terms of the biological and transmission processes that generate them. Rather than fitting a statistical distribution directly to observed delays, these models describe how pathogen replication, host response, infectiousness, and transmission interact to determine when key events occur. Their parameters therefore have direct biological interpretations, making it possible to distinguish competing explanations for observed transmission patterns.

This approach has already proved valuable in several settings. Pathogen-growth models have improved inference for incubation periods while retaining biologically interpretable parameters [11]. More recently, joint models of incubation periods have combined viral-load, viral-culture, symptom, and transmission data to infer otherwise unobservable infectiousness profiles [12], while mechanistic household transmission models have shown that assumptions about within-host biology and transmission can substantially influence inferred generation intervals [13]. Collectively, these studies illustrate the value of incorporating biological mechanisms into epidemiological inference, but they typically focus on individual delays or transmission processes rather than linking multiple epidemiological delays within a single framework.

Our objective is to model epidemiological delays as coupled outcomes of a common biological process rather than as separate statistical quantities. We therefore develop a unified event-time framework in which these four key epidemiological delays all arise from a shared within-host pathogen-growth model. The events are represented as stochastic events, in an approach related to the hazard-based formulation used in contact-interval models [14], but here transmission is linked to the same within-host process that governs the other event times to produce mutually consistent delay distributions [1, 2].

The proposed framework is applicable whenever infection, infectiousness, symptom onset, and transmission can be represented as linked biological event times. We focus here on acute person-to-person infections where pathogen load rises and falls over a relatively short time scale and has been shown to be associated with these biological events [15–17]. Under these conditions, pathogen growth provides a biologically plausible mechanism linking the major epidemiological delays [15–17]. Although we use a simple pathogen-growth model for illustration, the underlying event-time framework is flexible and can accommodate alternative models appropriate to other transmission settings. Importantly, the framework can be fitted to transmission-pair or serial-interval data alone, or jointly with additional biological data when available. To account for incomplete observation of event times, we incorporate an observation model that accommodates censored and partially observed infection and symptom-onset times. In this paper, we derive the resulting delay distributions, evaluate the framework using simulation studies, and demonstrate its application using SARS-CoV-2 and mpox transmission-pair data.

## 2 Methods

We represent the four key events as linked event times within an infector–infectee pair. We first define the relationships among these events, then specify a within-host pathogen-growth model and corresponding event hazards. We use these components to derive the latent-period, incubation-period, generation-interval, and serial-interval distributions before evaluating the framework through simulation and empirical applications.

### 2.1 Event-time representation of transmission

We represent transmission using a pair of individuals consisting of an infector and an infectee. The framework distinguishes underlying biological events from the epidemiological delays they generate. Figure 1 illustrates these relationships and establishes the notation used throughout the paper. These definitions follow the standard epidemiological framework for generation intervals and serial intervals [1, 5].

**Figure 1:**
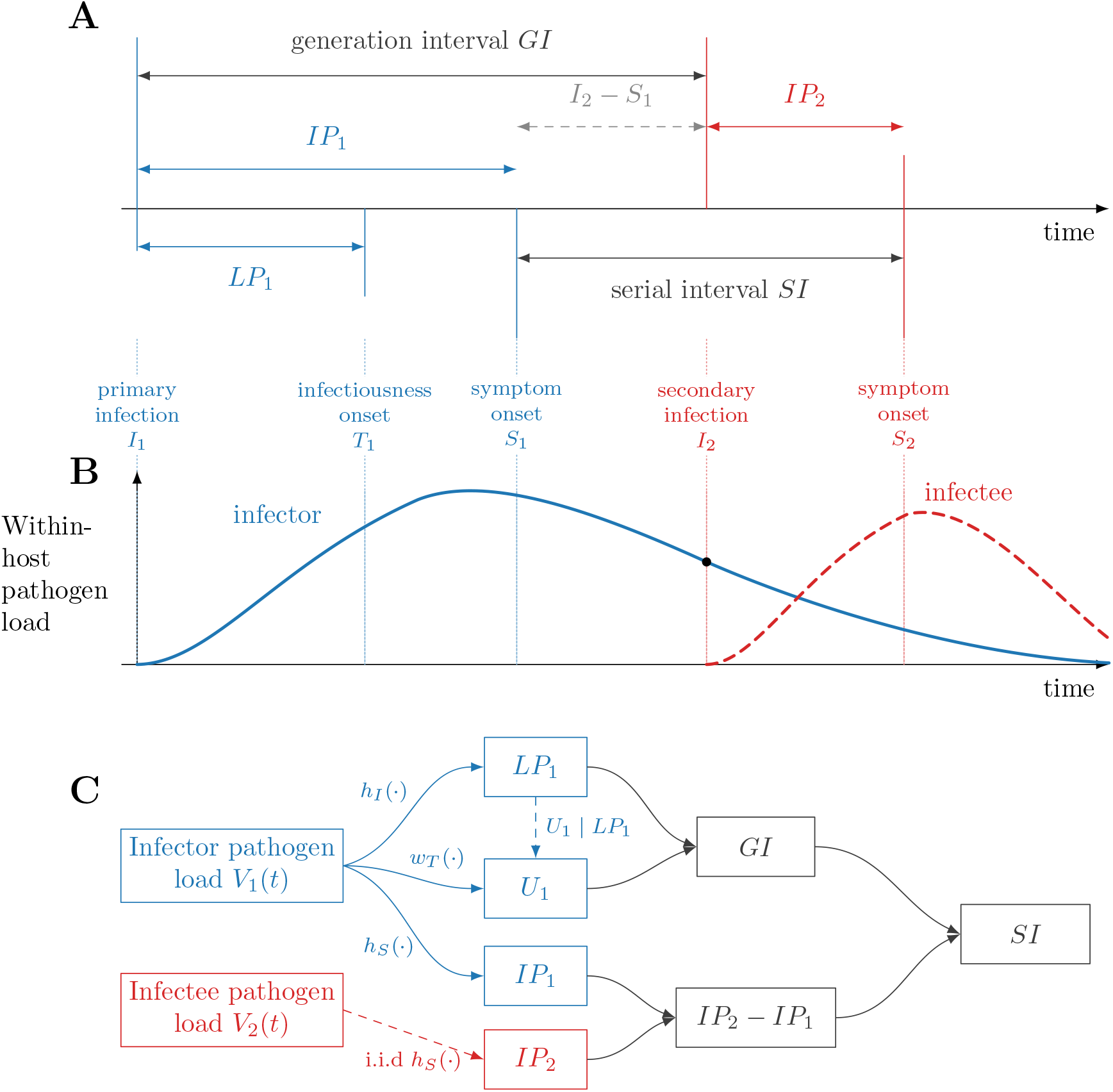
Epidemiological delays and their relationship to within-host pathogen dynamics. (A) Event times and epidemiological delays for an infector-infectee pair. Blue denotes events and intervals for the infector, red denotes those for the infectee, and gray denotes intervals spanning both individuals. The latent period and incubation period of the infector are *LP*_1_ = *T*_1_ − *I*_1_ and *IP*_1_ = *S*_1_ − *I*_1_, respectively; the generation interval is *GI* = *I*_2_ − *I*_1_ and the serial interval is *SI* = *S*_2_ − *S*_1_. The incubation period of the infectee is *IP*_2_ = *S*_2_ − *I*_2_. Transmission before symptom onset in the infector can produce negative values of *I*_2_ − *S*_1_ and of the serial interval. (B) Schematic pathogen-load trajectories associated with the event times. The infector trajectory governs infectiousness onset, symptom onset, and transmission at *I*_2_; infection of the secondary case initiates a separate pathogen-load trajectory that influences symptom onset at *S*_2_. (C) Dependency structure of the within-host event-time model. The infector pathogen-load trajectory *V*_1_(*t*) determines the latent period *LP*_1_, post-infectiousness transmission delay *U*_1_, and infector incubation period *IP*_1_ through a infectiousness-onset hazard *h*_*I*_(·), per-contact transmission weight *w*_*T*_ (·), and symptom-onset hazard *h*_*S*_(·), respectively. The infectee incubation period *IP*_2_ is generated analogously from the infectee trajectory *V*_2_(*t*) using the same symptom-onset model. The generation interval is *GI* = *LP*_1_ + *U*_1_, the difference in incubation periods is *X* = *IP*_2_ − *IP*_1_, and the serial interval is *SI* = *GI* + *X*. Dependence among *LP*_1_, *U*_1_, and *IP*_1_ arises through their shared dependence on *V*_1_(*t*); under the baseline assumptions, *IP*_2_ is conditionally independent of the infector-side event times. The framework links the epidemiological delays in panel A through the within-host processes shown in panel B, with panel C summarizing the resulting hierarchical dependence structure.

Consider a transmission pair consisting of an infector, indexed by 1, and an infectee, indexed by 2. Let *I*_1_ and *I*_2_ denote the infection times of the infector and infectee, respectively, *S*_1_ and *S*_2_ their symptom-onset times, and *T*_1_ the time at which the infector becomes infectious.

The latent period is the interval between infection of individual 1 and the onset of infectiousness,

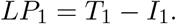

After becoming infectious, transmission occurs after a delay *U*_1_, giving the infection time of the secondary case,

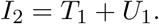

The generation interval is therefore

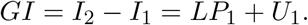

The incubation periods for the infector and infectee are

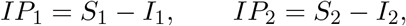

and the serial interval is

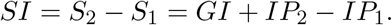

### 2.2 Within-host event-time model

The underlying event-time structure is fixed, but its component processes are modular and can be replaced or extended without changing the overall framework. Here, we consider a simple implementation in which the within-host pathogen-load trajectory drives the event hazards, providing a biologically plausible description of many acute viral infections [15–17].

Each event is represented by a hazard function describing its instantaneous rate over time. Alternative assumptions therefore correspond to different hazard functions while leaving the underlying event-time framework unchanged. Table 1 summarizes the parameters governing the within-host-informed event-time framework within this basic implementation, with specific components derived in the following subsections.

**Table 1:**
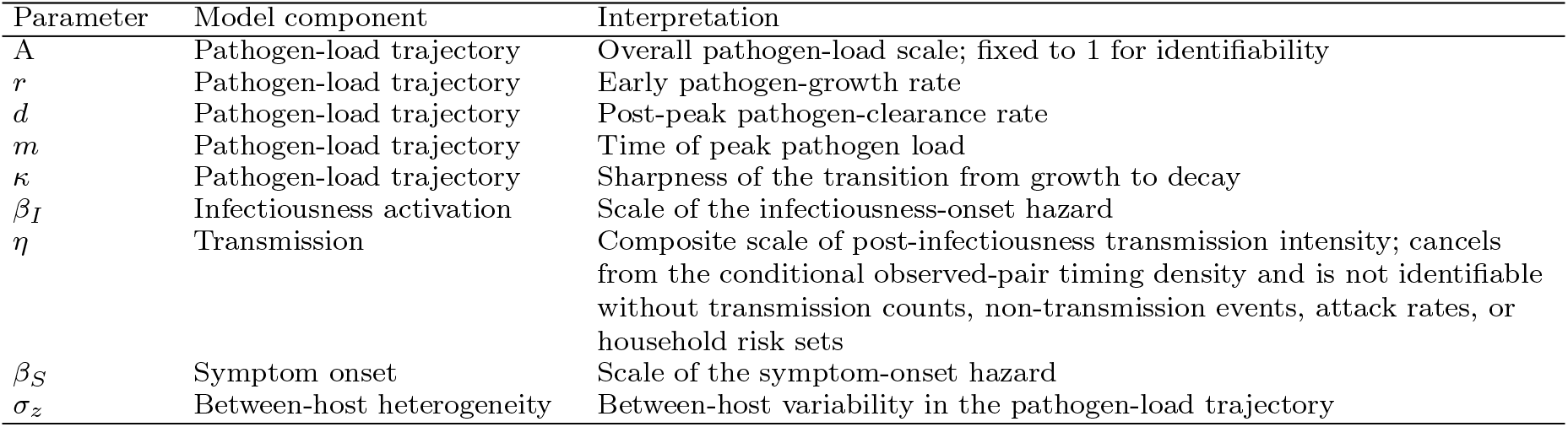
Parameters of the within-host-informed event-time framework in its basic implementation. The pathogen-growth parameters define the within-host trajectory, while the remaining parameters determine the hazards governing the four key biological events.

| Parameter | Model component | Interpretation |
| --- | --- | --- |
| $A$ | Pathogen-load trajectory | Overall pathogen-load scale; fixed to 1 for identifiability |
| $r$ | Pathogen-load trajectory | Early pathogen-growth rate |
| $d$ | Pathogen-load trajectory | Post-peak pathogen-clearance rate |
| $m$ | Pathogen-load trajectory | Time of peak pathogen load |
| $\kappa$ | Pathogen-load trajectory | Sharpness of the transition from growth to decay |
| $\beta_I$ | Infectiousness activation | Scale of the infectiousness-onset hazard |
| $\eta$ | Transmission | Composite scale of post-infectiousness transmission intensity; cancels from the conditional observed-pair timing density and is not identifiable without transmission counts, non-transmission events, attack rates, or household risk sets |
| $\beta_S$ | Symptom onset | Scale of the symptom-onset hazard |
| $\sigma_z$ | Between-host heterogeneity | Between-host variability in the pathogen-load trajectory |

#### 2.2.1 Pathogen-load dynamics

We denote pathogen load at time *t* since infection by *V* (*t*). To capture the characteristic rise, peak, and subsequent decline in pathogen load observed for many acute viral infections [11, 15, 17, 18], we use the following parsimonious trajectory

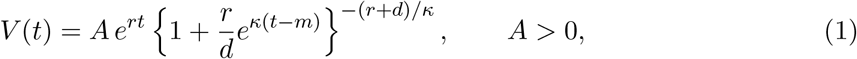

where *A >* 0 determines the overall pathogen-load scale, *r >* 0 is the early exponential growth rate, *d >* 0 is the post-peak exponential decay rate, *m* is the time of peak pathogen load, and *κ >* 0 controls the sharpness of the transition between growth and decay.

#### 2.2.2 Infectiousness onset

We assume that infectiousness becomes more likely as pathogen load increases, but that the pathogen-load threshold for infectiousness varies among individuals. We therefore represent infectiousness onset as a stochastic event whose instantaneous rate depends on the evolving pathogen-load trajectory.

We represent infectiousness onset by the hazard

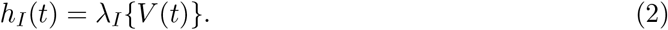

As a baseline specification, we assume the hazard is proportional to pathogen load,

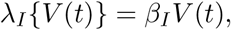

where *β*_*I*_ *>* 0 controls the scale of infectiousness activation.

#### 2.2.3 Transmission after infectiousness onset

Once an individual becomes infectious, transmission depends on both biology and opportunity. Even highly infectious individuals cannot transmit infection without contact with susceptible individuals, while frequent contacts may not result in transmission if pathogen load is low. We therefore model transmission as the combination of a contact process and a pathogen-load-dependent probability of successful transmission.

Conditional on infectiousness beginning at time *LP*_1_ = *l*, let *c*(*t*) denote the rate of transmission-relevant contacts at time *t* since infection, and let *w*_*T*_ (*t*) denote a non-negative per-contact transmission weight at time *t*. The transmission intensity at delay *U*_1_ after infectiousness onset is

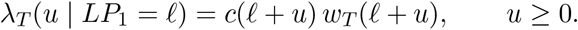

This formulation is closely related to pairwise contact-interval models [14], but here the transmission intensity is linked explicitly to the infector’s evolving pathogen-load trajectory and is used to describe the relative timing of transmission conditional on an observed transmission event.

The model therefore describes the timing of observed transmission events, conditional on transmission occurring, rather than the probability that transmission occurs within a complete infectious–susceptible risk set. In settings where household risk sets, repeated exposures, non-transmission events, or susceptible depletion are observed or can be modeled, this event-time component could instead be embedded within a fuller transmission model that accounts for these processes explicitly.

As a baseline specification, we assume a constant contact rate,

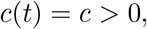

and take the per-contact transmission weight to be proportional to pathogen load,

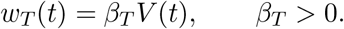

Here, *β*_*T*_ is a positive scaling parameter linking pathogen load to transmission intensity. Because the model conditions on the timing of an observed transmission event, the absolute scale of *w*_*T*_ (*t*) is not separately identified and cancels from the corresponding conditional timing distribution.

More generally, the per-contact transmission weight could be replaced by a bounded or saturating function of pathogen load without changing the overall framework. If such a function is constrained to lie in [0, 1], it can be interpreted directly as the probability that a transmission-relevant contact results in infection.

Under these assumptions, the conditional transmission intensity becomes

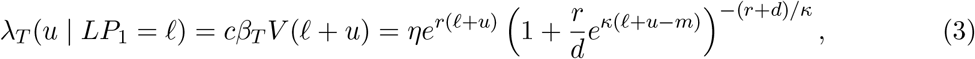

where *η* = *cβ*_*T*_ *A* and *u* ≥ 0.

#### 2.2.4 Symptom onset

Symptom onset is modeled analogously to infectiousness onset. As pathogen load increases, symptoms become more likely to develop, but individuals are not expected to develop symptoms at an identical pathogen-load threshold because symptom onset also depends on host responses and clinical variability [11]. We therefore represent symptom onset as a stochastic event whose instantaneous rate depends on the evolving pathogen-load trajectory.

The symptom-onset hazard is

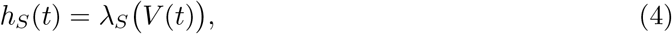

where *λ*_*S*_(·) maps pathogen load to the instantaneous rate of symptom onset.

Throughout this paper, we use a proportional hazard as the baseline specification,

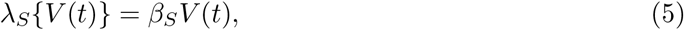

where *β*_*S*_ *>* 0 controls the rate at which symptoms emerge as pathogen load increases. Larger values of *β*_*S*_ correspond to earlier symptom onset for the same underlying pathogen-growth trajectory.

The overall pathogen-load scale *A* is not separately identifiable from the scale parameters linking pathogen load to infectiousness onset, symptom onset, and transmission. We therefore fix *A* = 1 throughout. Under this normalization, *β*_*I*_ and *β*_*S*_ are interpreted as hazard-scale parameters relative to the normalized pathogen-load trajectory, while the transmission scale is absorbed into the composite parameter *η*.

#### 2.2.5 Between-host heterogeneity

We represent between-host heterogeneity using an individual-level multiplicative random effect. For individual *i*,

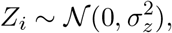

where *σ*_*z*_ *>* 0 controls the magnitude of between-host variability. Conditional on *Z*_*i*_ = *z*_*i*_, the infectiousness and symptom-onset hazards are

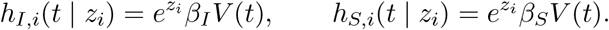

Thus, the same host-level random effect is shared by infectiousness onset and symptom onset within an individual, inducing dependence between the corresponding latent and incubation periods. The infector and infectee receive independent random effects,

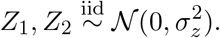

For the observed-pair transmission-delay distribution, the multiplicative factor 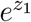 cancels when the transmission intensity is normalized over time, so the random effect influences transmission timing indirectly through the realized latent period rather than through an additional multiplicative transmission-scale term.

### 2.3 Epidemiological delay distributions

The event-time model introduced in Section 2.2 determines the latent period, generation interval, incubation period, and serial interval distributions derived below.

#### 2.3.1 Latent period

The latent period is the time from infection until the onset of infectiousness, conditional on infectiousness onset occurring. Because the total accumulated infectiousness-onset hazard may be finite under the pathogen-load trajectory used here, we normalize the event-time distribution by the probability that infectiousness onset occurs. Thus,

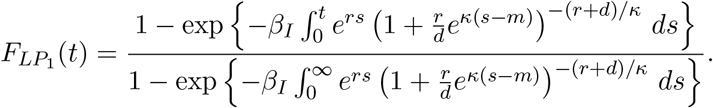

#### 2.3.2 Transmission delay and generation interval

The transmission delay, *U*_1_, is the interval between the onset of infectiousness and transmission to a secondary case. Because the model is fitted to observed transmission pairs, its distribution describes the relative timing of transmission conditional on an observed transmission event. This is analogous to the conditioning used in pairwise contact-interval models [14].

Because the model considers observed transmission pairs, we model the timing of a selected transmission event conditional on the latent period, rather than the waiting time to the first successful transmission event. Specifically, the transmission intensity is normalized over time to define the conditional timing distribution; this formulation does not reweight the latent-period distribution by the probability of onward transmission. Thus,

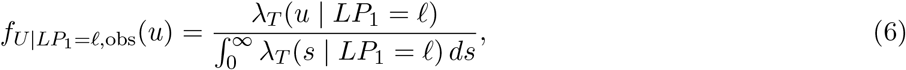

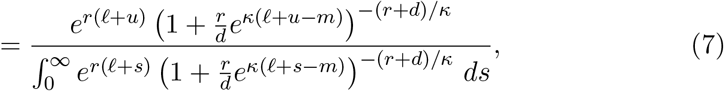

where *u* ≥ 0. The proportionality constant *η* cancels because conditioning on an observed transmission event depends only on the relative timing of transmission, not its absolute rate.

This conditional density describes the timing of observed transmission events rather than the waiting time to first successful transmission in a household or population risk set. Consequently, the probability of no onward transmission is not part of the likelihood and would instead be incorporated through models that explicitly represent risk sets, attack rates, offspring distributions, or non-transmission events.

The generation interval is obtained by combining the latent period and transmission delay. Its density is therefore

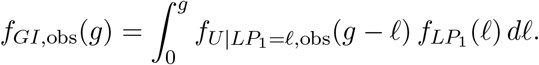

For numerical evaluation, this conditional density may be evaluated on a finite time grid and renormalized over the grid. Such a grid may be chosen to capture essentially all of the conditional probability mass, so this finite-grid normalization serves as a numerical approximation to the continuous conditional distribution above.

#### 2.3.3 Incubation period

The incubation period is derived analogously from the symptom-onset hazard and is defined conditional on symptom onset occurring. Under the proportional hazard in equation (5), the cumulative distribution is

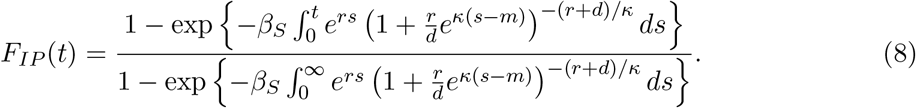

The corresponding conditional density is

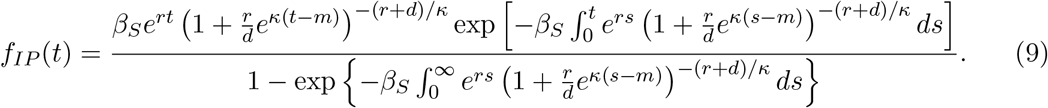

Thus, the incubation-period distribution is the symptom-onset event-time distribution conditional on symptom onset occurring.

For numerical evaluation, this conditional density may be evaluated on a finite time grid and renormalized over the grid. Grid bounds may be chosen so that residual conditional probability mass beyond the numerical support is negligible.

#### 2.3.4 Serial interval

The serial interval combines the generation interval with the difference between the incubation periods of the infectee and infector. Using the relationships introduced in Section 2.1,

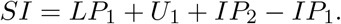

Many serial-interval models simplify this decomposition by assuming independence between the generation interval and the incubation periods [19–21]. In our framework, however, the infector-side quantities *LP*_1_, *U*_1_, and *IP*_1_ are dependent through their shared within-host pathogen-growth trajectory.

Let *z*_1_ denote the host-level random effect governing between-host variation in the infector’s pathogen-load trajectory, with density *p*(*z*_1_ | *σ*_*z*_), where *σ*_*z*_ *>* 0 controls the magnitude of between-host heterogeneity in the pathogen-load trajectory. Let *V*_1_(*t* | *z*_1_) denote the corresponding infector-specific trajectory. Conditional on *z*_1_, the latent period and incubation period are generated from their respective event-time distributions, while the post-infectiousness transmission delay is generated from the observed-pair transmission-timing distribution conditional on the realized latent period. The infector-side joint density is therefore

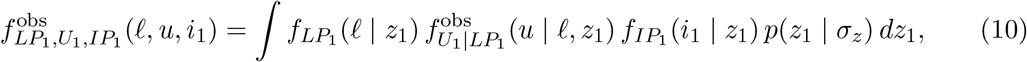

For

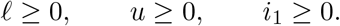

Thus, dependence among *LP*_1_, *U*_1_, and *IP*_1_ arises after marginalizing over the shared host-level random effect. Under the baseline model, the infectee has an independent host-level random effect and hence *IP*_2_ is independent of the infector-side event times conditional on the model parameters.

Let 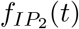 denote the resulting marginal incubation-period density for the infectee. The serial-interval density is then

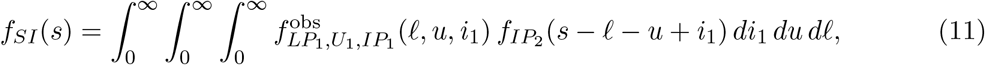

where 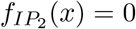 for *x <* 0.

#### 2.3.5 Connections to standard delay distributions

Simple choices of event hazards recover many of the statistical distributions commonly used to model epidemiological delays. A constant hazard gives an exponential waiting time, corresponding to an instantaneous event rate that does not change with time since infection. Allowing the hazard to increase or decrease as a power of time gives a Weibull distribution, so that the event becomes progressively more or less likely as infection progresses. An exponentially changing hazard gives a Gompertz distribution, allowing the timing of the event to become increasingly concentrated over the course of infection. Burr distributions can represent hazards that vary during the early course of infection but approach a constant at longer delays, allowing a pronounced mode while retaining an approximately exponential tail. An Erlang distribution arises when a delay is represented as the sum of a fixed number of independent exponential waiting times with a common transition rate. The Erlang distribution is the integer-shape special case of the gamma distribution, which more generally allows the shape parameter to take non-integer values. If the stage-specific transition rates differ, the resulting distribution is hypoexponential. Thus, commonly used delay distributions encode different assumptions about how the underlying timing process evolves over the course of infection. Further details are provided in Appendix A.

### 2.4 Model fitting and computation

All models were fitted in a Bayesian framework using Stan [22, 23] via the `cmdstanr` interface in R [24]. Sampling quality was assessed using 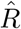, bulk and tail effective sample sizes, and Hamiltonian Monte Carlo diagnostics, including divergent transitions. We treated 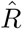 *<* 1.01 as indicating satisfactory convergence, while also inspecting effective sample sizes and divergent transitions. Model-specific priors, sampling settings, and diagnostic summaries are provided in Appendices C and D. Code implementing the models and analyses is publicly available at https://github.com/epiENGAGE/Within-host-time-delay-paper.

All event-time densities used in numerical convolution were evaluated on finite grids and normalized over their respective numerical supports. This grid normalization approximates the analytically defined distributions conditional on the corresponding event occurring; grid limits were selected to make truncation of the conditional tails negligible.

### 2.5 Simulation study design

We used four simulation studies to evaluate finite-sample recovery, performance across disease time scales, sensitivity to prior information, and joint recovery of multiple epidemiological delays. Full data-generating parameter values, prior specifications, numerical grids, and evaluation details are provided in Appendix D.

In the first simulation, we assessed finite-sample recovery under a well-specified SARS-CoV-2-like data-generating scenario with a mean generation interval of approximately 5 days. The priors were centered close to the data-generating parameter values, so this experiment evaluates recovery when the model and prior information are favorably specified rather than isolating information supplied by the serial-interval data alone. We simulated serial-interval datasets from the observed-pair within-host-informed model using the fixed parameter values given in Appendix D, which implied a generation-interval mean of 5.06 days and standard deviation of 2.43 days. For each sample size *n* ∈ {30, 50, 100}, we generated 100 replicate datasets from the model-implied serial-interval distribution and refitted the same model using the prior structure described in Appendix D.

In the second simulation, we evaluated recovery across different disease time scales. We constructed three data-generating regimes with model-implied mean generation intervals close to 3, 10, and 20 days, representing short, intermediate, and long-generation-interval settings. Each regime was generated using a distinct within-host parameter set, with the infectiousness-onset hazard scale numerically tuned to achieve the target mean generation interval. For each regime, we simulated *n* = 200 serial intervals from the corresponding observed-pair serial-interval distribution and refitted the model using broad common priors chosen to support all three regimes rather than favoring any single time scale.

In the third simulation, we assessed sensitivity to prior information and the learnability of latent within-host parameters when the likelihood was informed only by serial-interval observations, without jointly fitting pathogen-load or other biological data. We generated data using the intermediate-scale parameter set described in Appendix D. Each of 50 replicates comprised *n* = 200 training transmission pairs and 5,000 independent held-out transmission pairs, with the same training and held-out datasets used across prior scenarios within each replicate. We refitted each training dataset under a grid of prior scenarios varying both prior accuracy and prior strength. Prior means for the within-host trajectory parameters were centered on the true values, mildly misspecified, or strongly misspecified, with weak, moderate, or strong prior standard deviations. These perturbations were applied to (log *r*, log *d, m*, log *κ*, log *σ*_*z*_), while the infectiousness-activation and symptom-onset parameters retained broad baseline priors. This design represents settings in which external pathogen-load or shedding data provide varying amounts of information about the within-host trajectory.

In the fourth simulation, we assessed whether the framework could simultaneously recover multiple epidemiological delay distributions from serial-interval data under a well-specified event-time model and the specified prior structure. We used the same SARS-CoV-2-like data-generating parameter set as in the first simulation and generated 100 replicate datasets, each containing *n* = 150 serial intervals. For each replicate, we refitted the observed-pair within-host-informed model and derived posterior distributions for the latent-period, incubation-period, generation-interval, and serial-interval distributions. Recovery was assessed by comparing posterior estimates of the mean and standard deviation of each delay with the corresponding true model-implied values, and by comparing the fitted delay distributions with the known data-generating distributions. This simulation was designed to evaluate whether multiple epidemiological delays can be estimated jointly as linked outcomes of a common underlying event-time process under the assumed model.

Across the simulation studies, recovery of the generation-interval mean and standard deviation was assessed using bias and root mean squared error of the posterior median, the continuous ranked probability score (CRPS) for the full posterior distribution, empirical coverage of the 90% credible interval, and posterior interval width. For the prior-sensitivity analysis, we additionally assessed posterior predictive fit, held-out log predictive density, distance between the true and fitted serial-interval distributions, and whether posterior estimates of the within-host trajectory parameters moved closer to the known data-generating values relative to the prior means.

### 2.6 Empirical case studies

We applied the within-host-informed event-time framework to two empirical transmission-pair datasets. In both case studies, censoring and partial observation of infection and symptom-onset times were handled using the observation framework described in Appendix B. Predictive performance was compared using the Watanabe–Akaike information criterion (WAIC) [25] and approximate leave-one-out cross-validation information criterion (LOOIC) [26]. Both criteria estimate out-of-sample predictive performance, but WAIC uses a posterior variance-based correction for effective model complexity, whereas LOOIC approximates predictive accuracy by leaving out each observation in turn and evaluating its prediction from the remaining data.

#### 2.6.1 SARS-CoV-2

We applied the framework to serial-interval data from household transmission pairs observed during the replacement of the SARS-CoV-2 Delta variant by Omicron in the Netherlands [21, 27]. The aim was to assess whether the within-host-informed event-time model could recover generation-interval distributions consistent with the observed serial-interval data while providing a mechanistic interpretation of strain-level differences.

We compared two modeling approaches: a conventional lognormal generation-interval model [21], and the within-host-informed model in equation (11). For the within-host-informed model, weakly informative priors were specified on transformed parameters governing the pathogen-load trajectory and event-time processes. For the lognormal comparator, priors were placed directly on the generation-interval mean and standard deviation following [21]. We also fit these two models while developing likelihood models that capture the censored nature of such data (Appendix B). We compared the inferred generation-interval distributions, within-host parameters, posterior predictive fit, and predictive information criteria. Full prior specifications, computational settings, and model-comparison details are provided in Appendix C.1.

Finally, we used the inferred generation-interval distributions to estimate variant-specific reproduction numbers for Delta and Omicron. Because reproduction-number estimates can be sensitive to analytical choices, including the assumed generation-interval distribution [28], this provided a downstream comparison of the two generation-interval models. Following [21], we applied the renewal equation to smoothed variant-specific incidence curves. Because the model was fitted to observed household transmission pairs and does not include susceptible competition, depletion, or unobserved non-transmission events, we interpret the fitted distribution as an observed-pair, realized generation-interval distribution rather than an intrinsic generation-interval distribution for the underlying transmission process [1, 13]. For each variant, we estimated

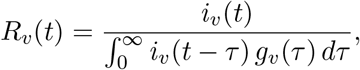

where *i*_*v*_(*t*) is the estimated incidence of infections with variant *v* at calendar time *t*, and *g*_*v*_(*τ*) is the fitted generation-interval density for that variant. The denominator represents the total infectiousness at time *t* contributed by individuals infected at earlier times, weighted by the probability of transmission *τ* days after infection.

#### 2.6.2 Mpox

As a secondary application, we applied the framework to mpox transmission-pair data to illustrate application beyond SARS-CoV-2 and examine the contribution of external viral-load information. We used publicly available data from [29], restricting the analysis to transmission pairs with a confidence score of 9 and high confidence in both the transmission link and symptom-onset dates. This yielded 34 transmission pairs, with serial intervals defined as the difference between symptom-onset dates of the infectee and infector.

We fitted three models. First, we fitted a conventional lognormal generation-interval model as a phenomenological comparator. In this model, the generation interval was specified directly as a lognormal distribution, and the serial-interval likelihood was obtained by combining the generation interval with a fixed incubation-period distribution. Second, we fitted the within-host-informed model using weak priors on the within-host trajectory parameters and event-hazard scales. Third, we robust-prior version of the within-host-informed model, in which external mpox viral-load data from [30] were used to construct informative priors for the within-host trajectory parameters. We refer to this as the Yang-informed prior.

The Yang-informed priors were constructed for

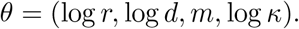

Because the viral-load measurements in Yang et al. are reported relative to symptom onset, whereas the event-time model parameterizes *V* (*t*) relative to infection, the posterior distribution for *m* from the viral-load model was shifted to the infection-time scale using an incubation-period offset.

We compared the lognormal model, the weak-prior within-host-informed model, and the Yang-informed robust-prior within-host-informed model. We also compared the Yang-informed prior with the weak-prior posterior for (log *r*, log *d, m*, log *κ*), to assess whether the within-host parameters favored by transmission-pair data alone were compatible with those implied by external viral-load measurements. Full specifications of the lognormal comparator, weak-prior within-host-informed model, and Yang-informed prior construction are provided in Appendix C.2.

## 3 Results

We first evaluate the framework in simulation before applying it to SARS-CoV-2 and mpox transmission-pair data.

### 3.1 Simulation study

The simulation study addressed four practical questions. First, how many transmission pairs are needed to recover generation-interval summaries? Second, does the framework remain accurate across diseases with different transmission time scales? Third, how sensitive is inference to prior information about the underlying within-host trajectory? Fourth, can the framework simultaneously recover multiple epidemiological delay distributions from serial-interval data alone?

#### 3.1.1 Small-sample recovery

Under this well-specified simulation setting, in which the priors were centered close to the data-generating parameter values, the within-host-informed event-time model recovered the mean and standard deviation of the generation-interval distribution across sample sizes of 30, 50, and 100 transmission pairs (Figure 2). At the smaller sample sizes, particularly *n* = 30, this recovery reflects information from both the simulated data and the favorably centered priors rather than the serial-interval data alone.

**Figure 2:**
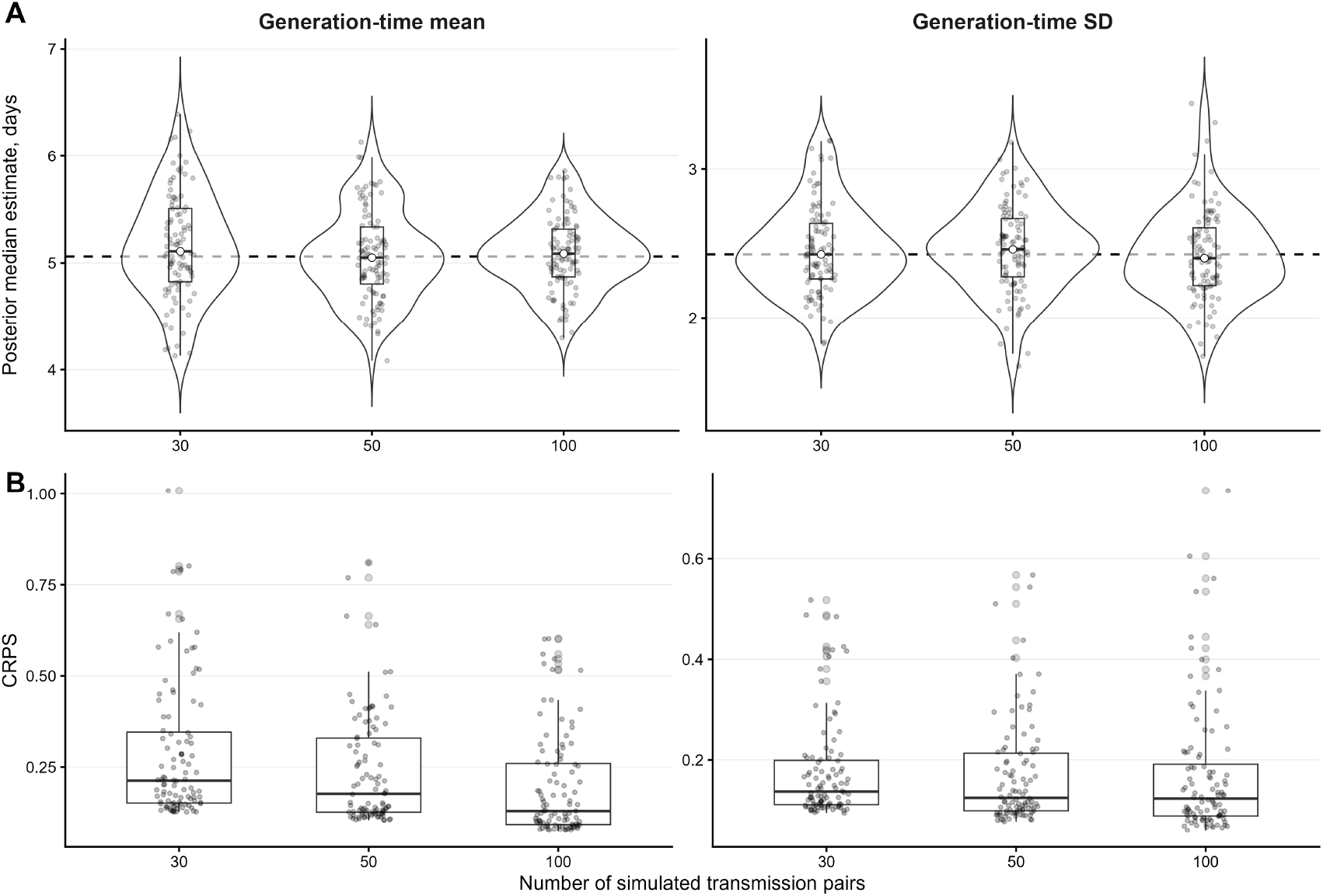
Recovery of generation-interval summaries from small transmission-pair datasets. Data were simulated from the within-host-informed observed-pair model using a fixed known parameter set and refitted at sample sizes of *n* = 30, 50, and 100 transmission pairs (100 simulation replicates per sample size). (A) Posterior median estimates of the generation-interval mean and standard deviation; dashed horizontal lines indicate the true data-generating values. (B) Continuous ranked probability score (CRPS) for the posterior distributions of the generation-interval mean and standard deviation. Lower CRPS indicates better probabilistic recovery of the true value.

The true generation-interval mean was 5.06 days. Across *n* = 30, 50, and 100, median bias was 0.05, −0.01, and 0.03 days, respectively, while RMSE decreased from 0.50 to 0.34 days and median absolute error from 0.32 to 0.21 days. Evaluation of the full posterior distribution showed the same pattern, with mean CRPS decreasing from 0.287 at *n* = 30 to 0.195 at *n* = 100. Empirical coverage of the nominal 90% posterior intervals was 94%, 94%, and 90%, while median interval width decreased from 1.93 to 1.13 days. Recovery of the generation-interval standard deviation was less sensitive to sample size, as mean CRPS changed only modestly from 0.176 to 0.169, although posterior intervals became progressively narrower.

Recovery of the generation-interval standard deviation was less sensitive to sample size. Mean CRPS changed from 0.176 at *n* = 30 to 0.169 at *n* = 100, while median 90% credible-interval width decreased from 1.54 to 1.01 days. Empirical coverage was 100%, 96%, and 91% for *n* = 30, 50, and 100, respectively.

#### 3.1.2 Stress-test across disease time scales

We used three contrasting generation-interval regimes as an illustrative stress test of model behavior across substantially different transmission time scales (Figure 3). The true mean generation intervals were 3.63, 9.99, and 20.00 days in the short, intermediate, and long generation-interval regimes, respectively. Median bias was 0.19, −0.36, and −0.10 days, while RMSE was 0.18, 0.49, and 0.98 days. Evaluation of the full posterior distributions showed increasing CRPS across the longer time scales, with mean CRPS values of 0.108, 0.296, and 0.535, respectively.

**Figure 3:**
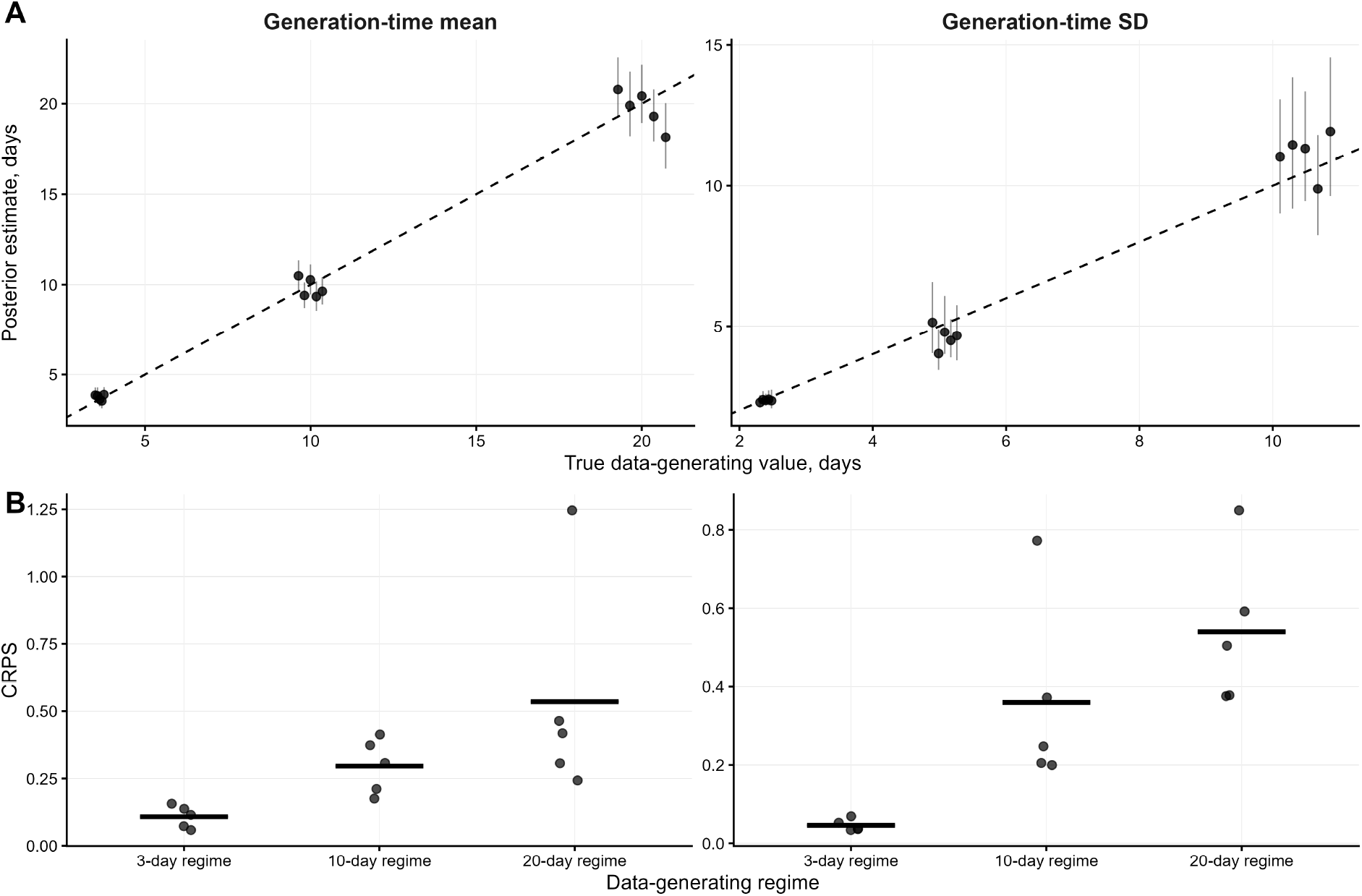
Recovery of generation-interval summaries across disease time scales. Data were simulated from three observed-pair within-host-informed models calibrated to mean generation intervals of approximately 3, 10, and 20 days, with *n* = 200 serial intervals per simulated dataset. (A) Posterior median estimates versus the true data-generating values; vertical bars denote 90% posterior intervals and the dashed diagonal line indicates perfect recovery. (B) Continuous ranked probability score (CRPS) for the posterior distributions of the generation-interval mean and standard deviation across the three data-generating regimes.

Median width of the 90% credible intervals increased from 0.83 to 3.34 days across the three regimes, and all five replicate credible intervals covered the true mean in each regime; however, with only five replicates per regime, these results should be interpreted descriptively rather than as estimates of frequentist coverage.

The generation-interval standard deviation was also recovered reasonably closely in these illustrative regimes, although uncertainty increased at longer time scales. Median bias was −0.04, −0.42, and 0.83 days, while RMSE was 0.06, 0.59, and 0.93 days. Mean CRPS was 0.046, 0.359, and 0.540 across the short, intermediate, and long-generation-interval regimes, respectively. Median width of the 90% credible intervals increased from 0.53 to 4.05 days. The true value was contained in 5/5, 4/5, and 5/5 replicate intervals in the short, intermediate, and long-generation-interval regimes, respectively. Given the small number of replicates, these proportions are reported descriptively and should not be interpreted as reliable estimates of interval coverage.

Taken together, these illustrative regimes suggest that the framework can recover generation-interval summaries across substantially different transmission time scales under the scenarios examined here. Precision decreased as the generation-interval scale increased, as reflected by larger CRPS values and wider posterior intervals. Because only five replicates were used per regime, this analysis is intended as a stress-test illustration rather than a formal Monte Carlo assessment of recovery.

#### 3.1.3 Prior sensitivity and parameter learnability

The third simulation examined the sensitivity of the within-host-informed model to prior information. By varying both prior accuracy and prior strength, we evaluated the benefits of informative mechanistic priors, the effects of prior misspecification, and the extent to which serial-interval data could update the within-host parameters away from misspecified prior values (Figure 4).

**Figure 4:**
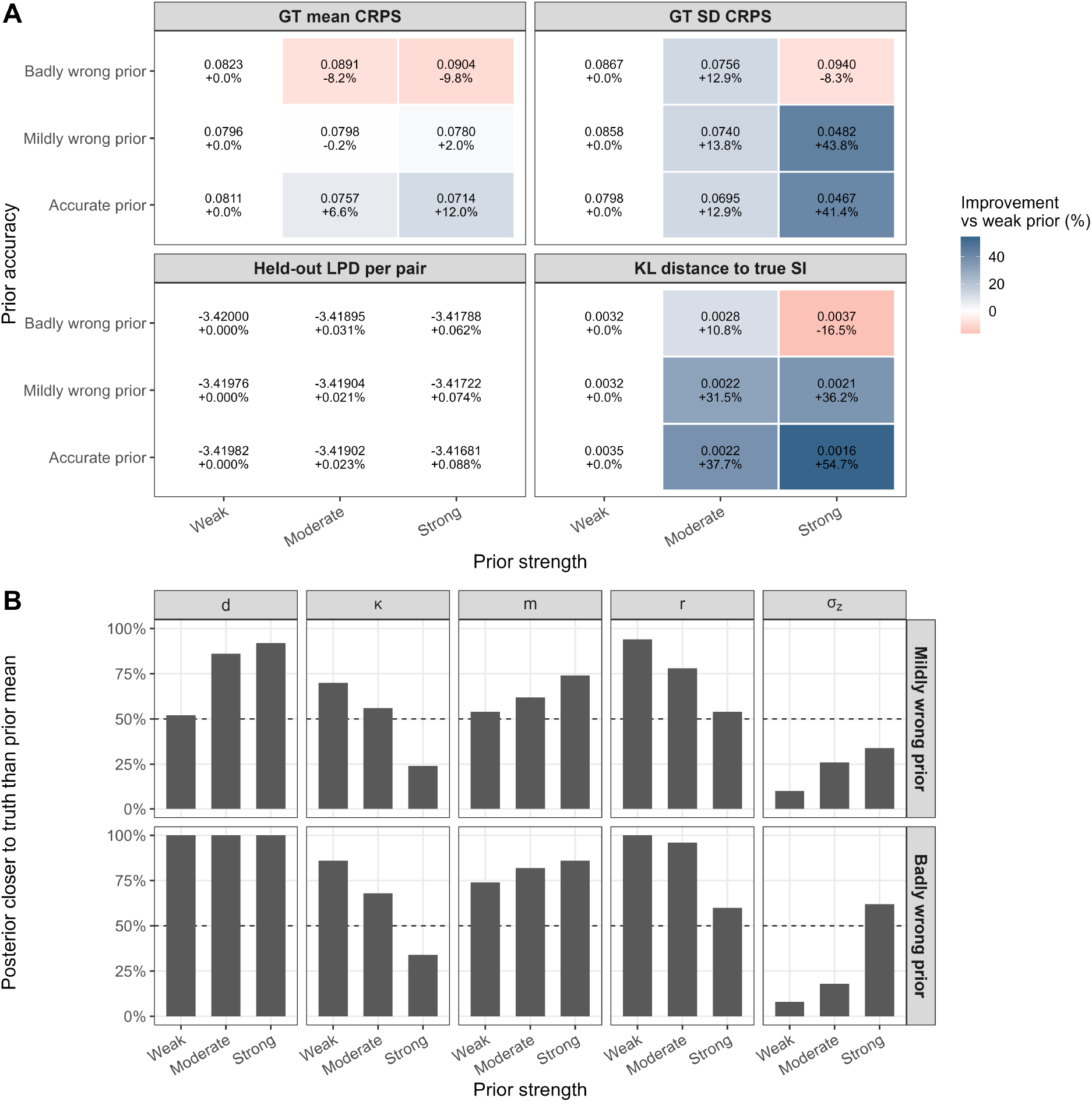
Figure 4: Prior sensitivity and parameter learnability. (A) Recovery of generation-interval summaries, recovery of the serial-interval distribution, and held-out predictive performance across combinations of prior accuracy and prior strength. Values beneath each tile show improvement relative to the weak prior within the same prior-accuracy class. Positive values indicate lower CRPS for the generation-interval mean and standard deviation, lower KL distance to the true serial-interval distribution, and higher held-out log predictive density. (B) Proportion of simulation replicates in which the posterior median moved closer to the true parameter value than the prior mean. Results are shown only for misspecified-prior scenarios. The dashed line marks 50%. Serial-interval data consistently corrected misspecified priors for *r, d*, and *m*, whereas *κ* and *σ*_*z*_ were less strongly learned from the serial-interval data.

When priors were accurately centered on the data-generating parameters, stronger prior information improved recovery of the generation-interval distribution and brought the fitted serial-interval distribution closer to the truth. Mild prior misspecification led to smaller changes. Strong mildly wrong priors gave little improvement in the generation-interval mean, but improved recovery of the generation-interval standard deviation and reduced the KL distance to the true serial-interval distribution. These results indicate that modest errors in prior location need not substantially degrade inference, particularly for broad distributional summaries. In contrast, strongly informative and misspecified priors degraded recovery of the latent generation-interval distribution. Moderate prior information could still be beneficial, but strong misspecification led to prior domination and poorer inference.

Held-out log predictive density (LPD) changed only weakly across scenarios. Notably, the strong badly wrong prior slightly improved held-out predictive density relative to the weak prior, despite worsening recovery of the generation-interval mean and standard deviation. Thus, good predictive performance for the observed serial-interval distribution does not necessarily imply accurate recovery of the underlying generation-interval distribution or within-host parameters.

Figure 4B examines parameter learnability. The extent to which posterior estimates moved toward the true parameter values varied across parameters. For the growth rate *r*, clearance rate *d*, and peak time *m*, the posterior frequently moved closer to the true values, particularly under weak or moderate misspecified priors. This suggests that serial-interval data contain some information about the broad timing of the within-host trajectory. Posterior estimates moved less consistently toward the true values for the transition-sharpness parameter *κ* and the between-host heterogeneity parameter *σ*_*z*_, especially under strongly misspecified priors, suggesting that these parameters were less strongly learned from serial-interval data alone and remained more influenced by the prior.

#### 3.1.4 Joint recovery of epidemiological delays

The fourth simulation assessed whether the framework could recover several epidemiological delay distributions simultaneously under the assumed event-time model. The model recovered the latent-period, incubation-period, generation-interval, and serial-interval distributions closely across the 100 simulated datasets (Figure 5). The median fitted density closely followed the true data-generating density for all four delays, while uncertainty across simulation replicates was greatest for the latent- and incubation-period distributions and narrower for the generation-interval and serial-interval distributions.

**Figure 5:**
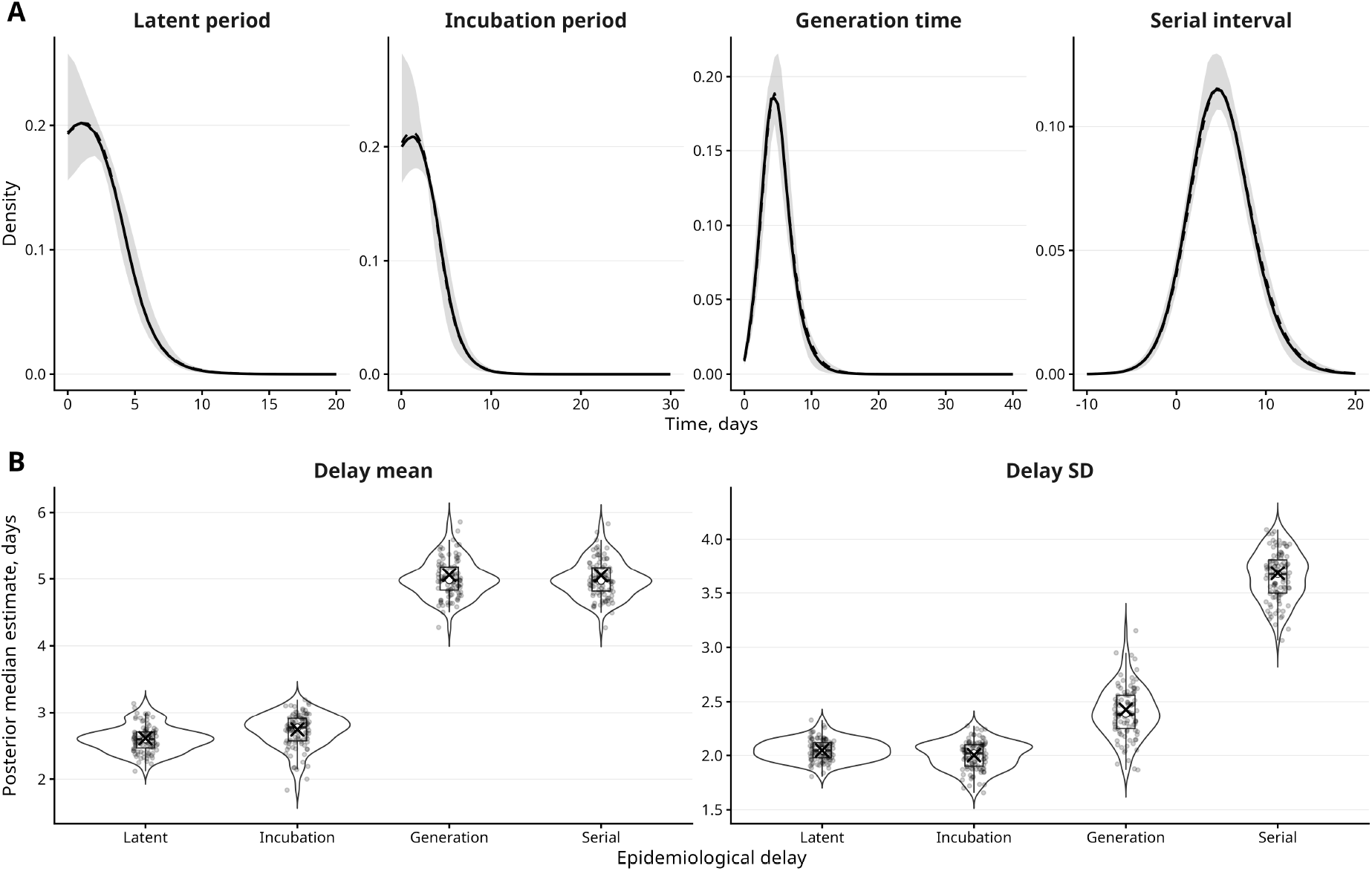
Joint recovery of epidemiological delay distributions from serial-interval data. One hundred datasets of *n* = 150 serial intervals were simulated from the within-host-informed observed-pair model and independently refitted. (A) Recovery of the complete latent-period, incubation-period, generation-interval, and serial-interval distributions. Dashed lines show the true data-generating densities, solid lines show the median fitted density across simulation replicates, and shaded regions show the 5th–95th percentile range of fitted densities across replicates. (B) Recovery of the mean and standard deviation of each delay distribution. Violin plots show the distribution of posterior median estimates across simulation replicates, boxplots indicate interquartile ranges, points show individual replicates, and crosses indicate the true data-generating values.

Recovery of the delay means was accurate across all four epidemiological quantities (Table 2). The median biases of the latent-period and incubation-period means were −0.02 and 0.03 days. The corresponding RMSEs were 0.20 and 0.26 days. The median bias of the generation-interval and serial-interval means were both −0.08 days. RMSEs were 0.28 days for the generation-interval mean and 0.28 days for the serial-interval mean. Mean CRPS values were similar across the four delay means, ranging from 0.159 for the serial interval to 0.171 for the incubation period.

**Table 2:**
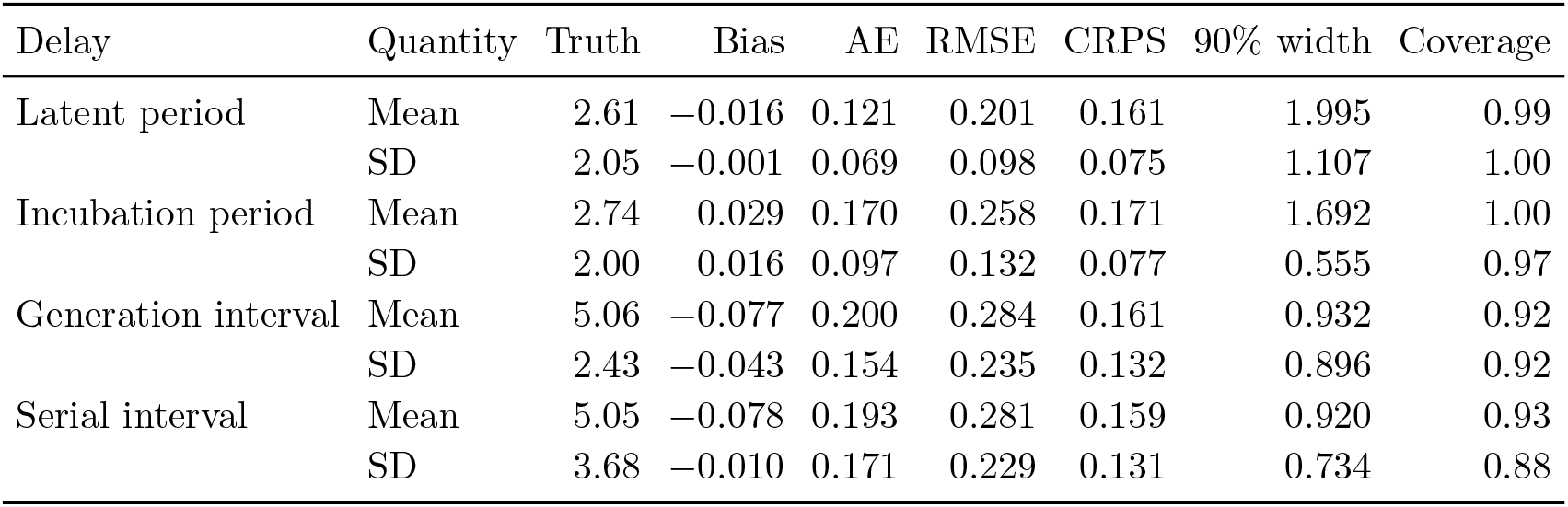
Recovery of jointly inferred epidemiological delay summaries. Results summarize 100 simulation replicates with *n* = 150 serial intervals per replicate. Bias and absolute error refer to the posterior median relative to the known data-generating value. RMSE is calculated across posterior medians, CRPS evaluates the full posterior distribution of each scalar summary, and coverage refers to the central 90% posterior interval.

The standard deviations of the four delay distributions were recovered with little systematic bias. Median bias was less than 0.05 days for every standard-deviation target. RMSE ranged from 0.10 days for the latent-period standard deviation to 0.24 days for the generation-interval standard deviation, while mean CRPS ranged from 0.075 to 0.132.

Posterior interval calibration differed somewhat among the delays. Coverage of the 90% intervals was 92–93% for the generation-interval mean and standard deviation and the serial-interval mean, and 88% for the serial-interval standard deviation. In contrast, intervals for the latent- and incubation-period summaries were more conservative, with empirical coverage between 97% and 100%. This greater uncertainty was also visible in Figure 5A, particularly for the latent and incubation-period densities.

The joint recovery also preserved differences among the epidemiological delays implied by the common event-time structure. In particular, the true generation-interval and serial-interval means were nearly identical (5.06 and 5.05 days), whereas their standard deviations differed substantially (2.43 versus 3.68 days). The model recovered both features simultaneously. Thus, recovery was not limited to reproducing a common overall transmission time scale; the framework recovered distinct latent, symptom-onset, transmission, and symptom-to-symptom timing distributions and the relationships among them from a single fitted model.

### 3.2 SARS-CoV-2 Application

We applied the within-host-informed event-time framework to household transmission-pair data collected during replacement of the SARS-CoV-2 Delta variant by Omicron in the Netherlands [21, 27]. Transmission pairs were stratified according to S-gene target failure (SGTF), which served as a proxy for Omicron during this period, while non-SGTF cases were predominantly associated with Delta infection [21, 27]. Following [21], a single infectee was randomly selected for each infector to reduce dependence among observations. After restricting the analysis to within-household transmission pairs, the final dataset comprised 644 Omicron (SGTF) and 1,333 Delta (non-SGTF) transmission pairs.

#### 3.2.1 Predictive performance and fit to observed serial intervals

The within-host-informed event-time model provided better predictive performance than the conventional lognormal model for both Omicron and Delta transmission pairs under the uncensored formulation (Table 3). For Omicron, the within-host-informed model improved both WAIC and LOOIC by 30.6 points relative to the lognormal model, with an SE of 10.6 for the paired LOOIC difference. For Delta, the corresponding improvement was 45.2 LOOIC points (SE 11.6). In both cases, the estimated improvement was several times larger than its standard error, indicating that the predictive advantage of the within-host-informed model was large relative to the uncertainty in the model comparison. The within-host-informed model had a larger effective number of parameters (*p*_WAIC_ ≈ 4–5 compared with approximately 1–2 for the lognormal model), but nevertheless achieved lower WAIC and LOOIC values, indicating better predictive performance after accounting for the additional model complexity.

**Table 3:** Predictive comparison of lognormal and within-host-informed (WH-informed) serial-interval models fitted separately to Omicron and Delta household transmission pairs under uncensored and censored observation models. WAIC values and effective numbers of parameters (*p*_WAIC_) are shown as WH-informed / lognormal. For leave-one-out cross-validation, ΔLOOIC is defined as LOOIC_Lognormal_ − LOOIC_WH_, so positive values indicate better expected out-of-sample predictive performance for the WH-informed model. The standard error of ΔLOOIC was calculated from the paired pointwise differences in ELPD-LOO between the two models.

| Strain | Observation | WAIC | $p_{\text{WAIC}}$ | $\Delta\text{LOOIC}$ (SE) |
| --- | --- | --- | --- | --- |
| Omicron | Uncensored | 2829.6 / 2860.2 | 4.53 / 1.02 | 30.6 (10.6) |
| Omicron | Censored | 2837.8 / 2948.3 | 3.09 / 0.69 | 110.6 (18.4) |
| Delta | Uncensored | 6333.8 / 6379.0 | 4.16 / 1.57 | 45.2 (11.6) |
| Delta | Censored | 6370.9 / 6372.1 | 3.51 / 1.63 | 1.2 (3.8) |

As a sensitivity analysis, we also fitted censored versions of both models to account for symptom-onset dates being recorded to the nearest day rather than observed continuously. Because the censored and uncensored formulations use different likelihoods, their WAIC and LOOIC values should not be compared directly. We therefore restricted model comparisons to models fitted under the same observation formulation. Under the censored formulation, the within-host-informed model reduced LOOIC by 110.6 points relative to the censored lognormal model for Omicron (SE 18.4); this difference was again large relative to its uncertainty, supporting a clear predictive advantage for the within-host-informed model. For Delta, by contrast, the two censored models differed by only 1.2 LOOIC points (SE 3.8), so the estimated difference was small relative to its uncertainty and the models could not be clearly distinguished in predictive performance.

PSIS-LOO diagnostics indicated no problematic observations, with all Pareto-*k* values below 0.5 for every fitted model. The maximum Pareto-*k* values for the WH-informed and lognormal models, respectively, were 0.35 and −0.01 for uncensored Omicron, 0.13 and 0.05 for uncensored Delta, 0.11 and 0.05 for censored Omicron, and 0.18 and 0.08 for censored Delta.

All models reproduced the observed serial-interval distributions for Omicron and Delta (Appendix E, Figure S1), with the clearest visual improvement for Omicron in the peak and tails of the distribution.

Across all fitted models, convergence diagnostics indicated satisfactory posterior sampling. The maximum 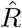 was 1.01, minimum bulk effective sample sizes exceeded 500, and minimum tail effective sample sizes exceeded 400 for both variants.

#### 3.2.2 Generation-interval distributions

All four models inferred shorter generation intervals for Omicron than Delta, although the estimated means varied (Figure 6). Under the lognormal model, the posterior median generation interval was 2.97 days (95% CrI: 2.73–3.22) for Omicron and 3.85 days (95% CrI: 3.71–3.99) for Delta. Under the within-host-informed model, the corresponding posterior medians were 3.12 days (95% CrI: 2.85–3.39) for Omicron and 3.99 days (95% CrI: 3.83–4.17) for Delta. Under the censored within-host-informed model, the posterior medians were 3.00 days (95% CrI: 2.82– 3.18) for Omicron and 3.76 days (95% CrI: 3.62–3.90) for Delta. Under the censored lognormal model, the posterior median generation interval was 3.12 days (95% CrI: 2.85–3.41) for Omicron and 3.81 days (95% CrI: 3.67–3.94) for Delta.

**Figure 6:**
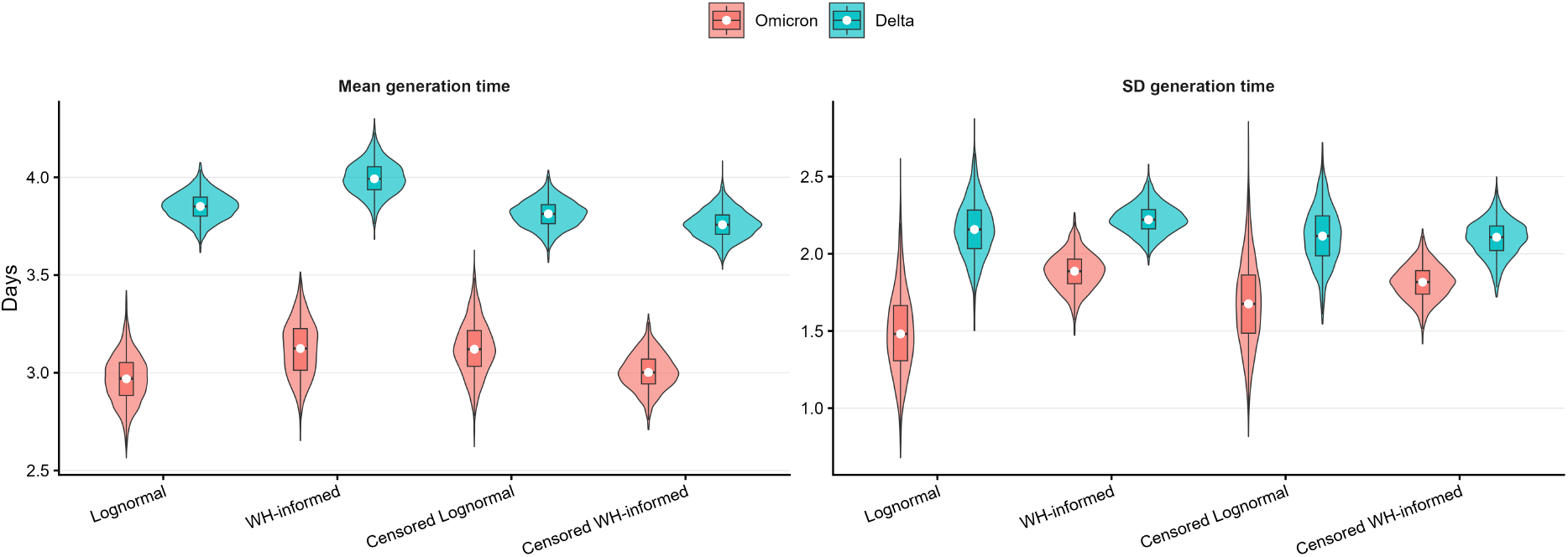
Posterior distributions of the generation-interval mean and standard deviation for Omicron and Delta under the lognormal, within-host-informed, and censored within-host-informed models. Violin plots show posterior draws, boxplots indicate interquartile ranges, and white points denote posterior medians.

The differences among the models reflect how variation is allocated between latent transmission timing and the observation process. The lognormal model estimates the generation-interval distribution directly with the serial-interval likelihood, whereas the within-host-informed model derives *GI* = *LP*_1_ + *U*_1_, linking both components through the shared pathogen-load-driven event-time framework.

This additional structure allows the model to redistribute variation among biologically linked latent events rather than requiring all features of the observed serial-interval distribution to be represented through the parameters of a single generation-interval distribution. The longer generation intervals inferred by the uncensored within-host-informed model are therefore consistent with the greater flexibility of the mechanistic decomposition. Further, explicitly accounting for day-level symptom-onset recording shifted the within-host-informed estimates downward relative to the lognormal for both variants. Importantly, the same censoring adjustment produced a smaller change in the lognormal estimates, suggesting that the remaining differences between model classes are not explained by observation error alone. Together with the substantially better predictive performance of the censored within-host-informed model for Omicron, these results suggest that the mechanistic framework is able to capture structure in the transmission-timing data that is not well represented by a simple lognormal generation-interval distribution, while for Delta the two censored formulations yield more similar estimates and predictive performance.

The models also differed in their inferred generation-interval variability. Under the lognormal model, the posterior standard deviation was lower for Omicron than for Delta and showed greater posterior uncertainty, particularly for Omicron. Under the within-host-informed models, posterior standard deviations were more similar across strains and concentrated around approximately 1.9–2.3 days.

#### 3.2.3 Comparison with external viral-kinetic estimates

To assess the biological plausibility of the inferred within-host trajectories, we compared the estimated trajectory parameters with independent viral-kinetic studies (Table 4). Because the model was fitted to transmission-pair timing data using weakly informative priors rather than to longitudinal viral-load measurements, agreement was evaluated primarily in terms of biologically plausible parameter ranges and relative differences between variants rather than exact numerical correspondence.

**Table 4:** Posterior estimates of the within-host trajectory parameters for Omicron and Delta compared to representative values from published SARS-CoV-2 viral-kinetic studies. Posterior estimates are medians with 95% credible intervals. The transformed quantities log(2)*/r* and log(100)*/d* represent the early exponential-growth doubling time and the model-implied time from peak pathogen load to 1% of the peak, respectively, under the post-peak approximation *V* (*t*) ∝ exp(−*dt*). All rates in the table are expressed on the natural-log scale.

| Quantity | Omicron | Delta | External comparator |
| --- | --- | --- | --- |
| $r$ | 0.46 [0.09, 1.92] d<sup>-1</sup> | 0.33 [0.08, 1.13] d<sup>-1</sup> | Comparable to the rising-phase exponential growth rate in log-scale viral-load models. Gunawardana et al. reported Omicron BA.1 viral-load doubling times of 3.3–3.5 h during the proliferation phase, equivalent to natural-log growth rates of approximately 4.75–5.04 d<sup>-1</sup> [31]. |
| $d$ | 0.73 [0.58, 0.87] d<sup>-1</sup> | 0.66 [0.53, 0.77] d<sup>-1</sup> | Comparable to the post-peak exponential decline rate in log-scale viral-load models. Jones et al. estimated approximately 0.39 d<sup>-1</sup>; Hay et al.’s estimates imply approximately 0.74 d<sup>-1</sup> for Omicron BA.1, 0.55 d<sup>-1</sup> for boosted BA.1, and 0.61 d<sup>-1</sup> for Delta [32, 33]. |
| $\log(100)/d$ | 6.29 [5.32, 7.95] d | 6.99 [5.95, 8.76] d | Model-implied time from peak to 1% of peak under $V(t) \propto \exp(-dt)$. Hay et al. reported Omicron BA.1 clearance durations of 6.2 d [5.8, 6.6] in individuals with one or two vaccine doses and 8.4 d [8.0, 8.7] in boosted individuals; they also reported a Delta clearance duration of 7.6 d [7.2, 8.0] in individuals with one or two vaccine doses [33]. |
| $m$ | 1.51 [0.81, 2.09] d | 2.08 [0.77, 3.06] d | Comparable to time from exposure to peak viral load. Russell et al. estimated peak Ct timing at 5.9 d [5.2, 6.7] after exposure for BA.1 and 6.6 d [5.1, 8.3] for Delta [18]. |
| $\kappa$ | 3.67 [1.41, 12.50] | 2.01 [0.73, 9.36] | No direct quantitative comparator. Piecewise-linear viral-load models impose an abrupt transition between proliferation and clearance, whereas $\kappa$ estimates the smoothness or sharpness of that transition. |

Agreement with external viral-kinetic studies varied considerably across parameters. The strongest quantitative agreement was observed for the post-peak decline parameter *d*, whereas the inferred peak timing *m* agreed primarily in the relative ordering of Omicron and Delta rather than in absolute timing. By contrast, the early growth parameter *r* and the transition-sharpness parameter *κ* were only weakly constrained by the transmission-pair data.

The early growth-rate parameter *r* should therefore be interpreted cautiously. The model inferred higher growth rates for Omicron than Delta, with posterior medians of *r* = 0.46 d^*−*1^ (95% CrI: 0.09–1.92) and *r* = 0.33 d^*−*1^ (95% CrI: 0.08–1.13), respectively. The corresponding model-implied doubling times were 1.49 days (95% CrI: 0.36–7.59) for Omicron and 2.10 days (95% CrI: 0.62–8.34) for Delta. Although the relative ordering is consistent with earlier within-host timing for Omicron, these proliferation rates remain substantially lower than estimates obtained directly from longitudinal viral-load data. For example, Gunawardana et al. reported BA.1 doubling times of approximately 3.3–3.5 h, corresponding to natural-log growth rates of approximately 4.75–5.04 d^*−*1^. This discrepancy indicates that serial-interval data provide information about the relative timing of early within-host dynamics but only weakly identify the absolute viral proliferation rate.

The post-peak decline parameter *d* showed close quantitative agreement with published viral-kinetic estimates. Posterior medians were *d* = 0.73 d^*−*1^ (95% CrI: 0.58–0.87) for Omicron and *d* = 0.66 d^*−*1^ (95% CrI: 0.53–0.77) for Delta. The Omicron estimate closely matches the decline scale implied by published BA.1 clearance estimates. In contrast, accounting for day-level symptom-onset recording shifted the Delta decline rate downward, corresponding to a longer inferred post-peak clearance period.

The corresponding peak-to-1% summaries support the same conclusion. The inferred post-peak decline corresponded to 6.29 days (95% CrI: 5.32–7.95) for Omicron and 6.99 days (95% CrI: 5.95–8.76) for Delta from peak pathogen load to 1% of peak, values that are broadly consistent with published estimates of SARS-CoV-2 clearance duration across different immune-history groups [33]. Although the model did not recover a pronounced difference in post-peak decline between Omicron and Delta, the inferred clearance time scales remained biologically plausible.

The inferred peak-time parameter *m* agreed with external evidence in relative ordering but not in absolute timing. The posterior medians were *m* = 1.51 days (95% CrI: 0.81–2.09) for Omicron and *m* = 2.08 days (95% CrI: 0.77–3.06) for Delta. This is consistent with Russell et al., who estimated earlier peak Ct timing for BA.1 than Delta [18]. However, the absolute peak times were substantially earlier than the approximately 6 days after exposure reported in longitudinal viral-load studies.

The transition-sharpness parameter *κ* was also only weakly constrained by the transmission-pair data. The posterior median was higher for Omicron than Delta (3.67 [95% CrI: 1.41–12.50] for Omicron and 2.01 [95% CrI: 0.73–9.36] for Delta), suggesting a sharper transition between pathogen growth and clearance for Omicron. However, most viral-kinetic studies do not estimate a directly comparable transition-sharpness parameter, so the absolute values of *κ* cannot be readily compared with external estimates.

### 3.3 Mpox application

We next applied the framework to 34 high-confidence mpox transmission pairs to assess its performance beyond SARS-CoV-2 and to examine the contribution of external viral-load information. Both within-host-informed models had lower LOOIC point estimates than the log-normal comparator (Table 5). The weak-prior within-host-informed model reduced LOOIC by 6.62 points relative to the lognormal model, with a paired standard error of 1.65. The Yang-informed robust-prior model reduced LOOIC by 2.44 points relative to the lognormal model, with a paired standard error of 2.23. Thus, the predictive improvement for the weak-prior model was large relative to its uncertainty, whereas the estimated improvement for the Yang-informed model was of similar magnitude to its standard error. Direct comparison of the two within-host-informed models gave a LOOIC difference of 4.18 points in favor of the weak-prior model, with a paired standard error of 2.44, indicating that the difference between the two formulations was modest relative to its uncertainty. PSIS-LOO diagnostics were satisfactory overall: the maximum Pareto-*k* values were 0.11 for the lognormal model, 0.28 for the weak-prior within-host-informed model, and 0.50 for the Yang-informed model. No observations had Pareto-*k >* 0.7 in any model; one observation in the Yang-informed model had Pareto-*k >* 0.5, whereas none exceeded 0.5 in the other two models.

**Table 5:** Predictive comparison of the lognormal and within-host-informed models fitted to the high-confidence mpox transmission-pair dataset. WAIC and *p*_WAIC_ are reported for each model. For the within-host-informed models, ΔLOOIC is defined as LOOIC_Lognormal_ − LOOIC_WH_, so positive values indicate better expected out-of-sample predictive performance than the log-normal comparator. Standard errors were calculated from the paired pointwise ELPD-LOO differences between models.

| Model | WAIC | $p_{\text{WAIC}}$ | $\Delta\text{LOOIC}$ (SE) |
| --- | --- | --- | --- |
| Lognormal | 220.67 | 1.42 | – |
| WHI, weak priors | 214.01 | 2.03 | 6.62 (1.65) |
| WHI, Yang-informed robust prior | 218.18 | 2.83 | 2.44 (2.23) |

The similar predictive performance of the two within-host-informed models does not imply the same within-host trajectory. To assess the mechanistic agreement, we compared the Yang-informed prior with the posterior obtained from under weak priors (Figure 7). This comparison evaluates whether the within-host trajectories supported by transmission-pair data alone are consistent with those implied by external viral-load measurements.

**Figure 7:**
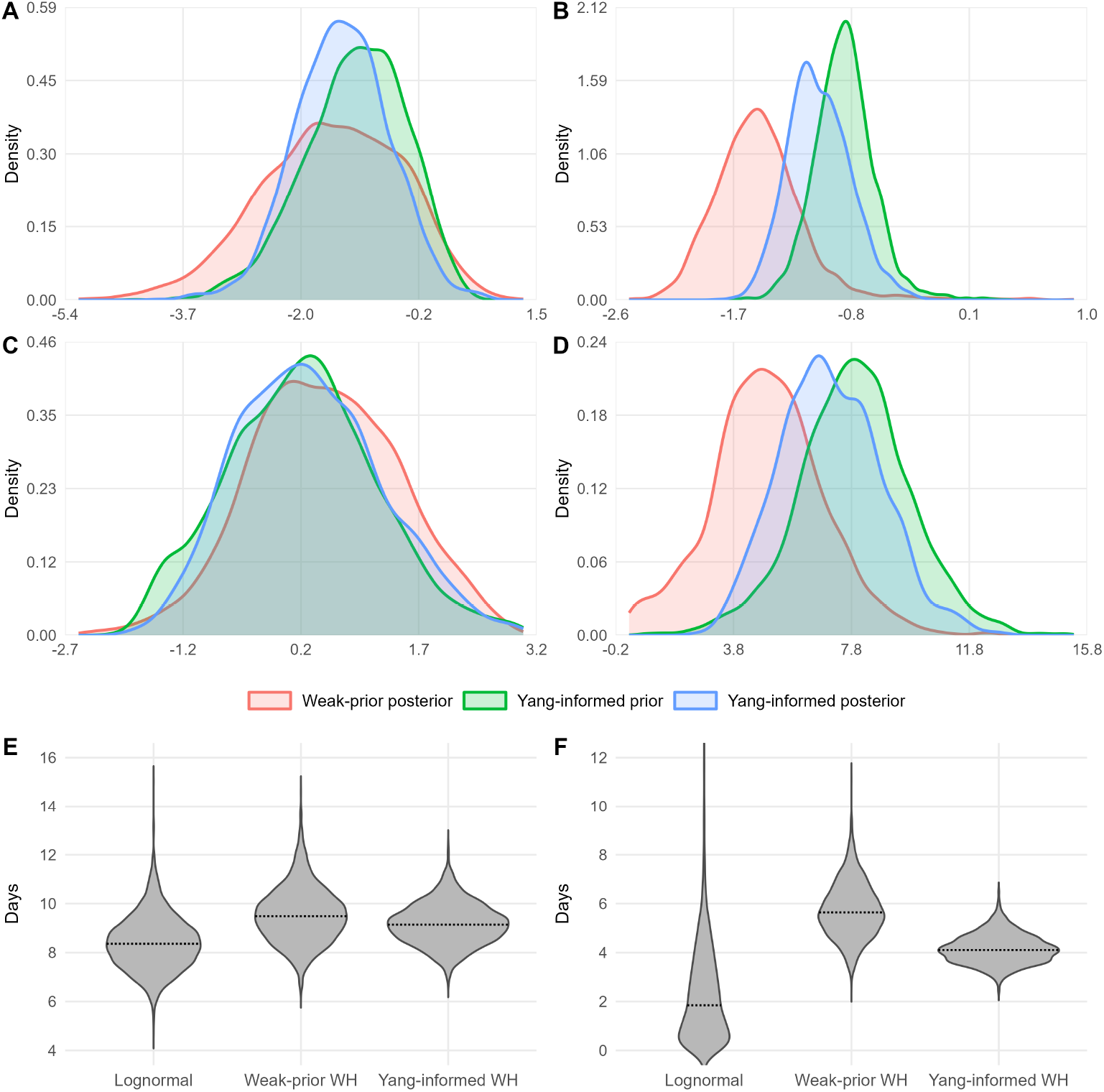
Comparison of models fitted to the mpox data. Panels A–D compare the weak-prior posterior, Yang-informed prior, and Yang-informed posterior for the within-host trajectory parameters: (A) log *r*, (B) log *d*, (C) log *κ*, and (D) *m*. The weak-prior posterior was obtained by fitting the within-host-informed model to the transmission-pair data using weakly informative priors, whereas the Yang-informed posterior incorporates informative priors derived from the external viral-load data. (E) and (F) show the posterior distributions of the generation-interval mean and standard deviation, respectively, for the weak-prior within-host-informed model, the Yang-informed within-host-informed model, and the lognormal comparator.

The weak-prior and Yang-informed analyses showed substantial overlap across all within-host trajectory parameters (Figure 7A–D), indicating broad agreement between the parameter values supported by the transmission-pair data and those implied by the external viral-load information. The distributions for log *r* and log *κ* overlapped strongly between the two analyses (Figure 7A,C).

Largest differences were observed for the decline parameter log *d* and the peak-time parameter *m* (Figure 7B). For log *d*, the weak-prior posterior was shifted toward lower values than the Yang-informed prior, implying slower post-peak decline under the transmission-pair data alone. The Yang-informed posterior shifted toward higher values, demonstrating that the external viral-load information substantially influenced inference on the decline phase of the within-host trajectory.

The peak-time parameter *m* showed a similar pattern (Figure 7D). Because the Yang viral-load data are reported relative to symptom onset rather than infection, they imply a later infection-time peak than the weak-prior transmission-pair analysis. Accordingly, the Yang-informed posterior shifted toward later values than the weak-prior posterior, indicating that external viral-load information also substantially influenced inference on peak timing.

The differences in log *d* and *m* are most informative when considered jointly. The weak-prior analysis favored an earlier peak and slower post-peak decline, whereas the Yang-informed analysis favored a later peak and faster decline. Despite these differences, both parameter combinations produced similar fits to the observed serial-interval data, demonstrating that the serial-interval likelihood does not uniquely identify the underlying within-host trajectory.

The lognormal model estimated a posterior median generation interval of 8.36 days (95% CrI: 6.50–10.86), compared with an increased 9.49 days (95% CrI: 7.54–12.09) under the weak-prior within-host-informed model and 9.15 days (95% CrI: 7.62–10.94) under the Yang-informed model (Figure 7E). Additionally, the posterior median standard deviation was 1.85 days (95% CrI: 0.06–6.66) under the lognormal model, compared with 5.64 days (95% CrI: 3.70–8.15) and 4.11 days (95% CrI: 3.10–5.44) under the weak-prior and Yang-informed within-host models, respectively (Figure 7F).

## 4 Discussion

We developed a within-host-informed event-time framework that jointly derives the latent period, incubation period, generation interval, and serial interval from a shared pathogen-growth process. By modeling the four key biological events as stochastic events linked through a common pathogen-growth trajectory, the framework distinguishes primitive biological events from derived epidemiological delays while making the assumptions connecting them explicit. The resulting delay distributions remain directly usable in conventional epidemiological analyses while retaining an explicit link between inferred transmission timing and the biological processes assumed to generate it. The shapes and relationships of these delay distributions therefore emerge from a common set of biologically motivated event hazards.

The simulation analyses showed that the framework could recover generation-interval characteristics across a range of settings. Generation-interval summaries were recovered from relatively small transmission-pair datasets, with uncertainty generally decreasing as sample size increased, and recovery remained good across short, intermediate, and long generation-interval regimes, although absolute uncertainty increased at longer time scales. The prior-sensitivity analysis further showed that serial-interval data alone were sufficient to recover generation-interval summaries more reliably than the underlying within-host parameters. Accurate mechanistic prior information could improve recovery, whereas strongly informative and substantially misspecified priors could degrade inference. Transmission-pair data therefore contain useful information about epidemiological delays but only partial information about their biological decomposition. In particular, parameters governing the detailed shape of pathogen growth and between-host heterogeneity were less strongly learned than the resulting generation-interval characteristics.

The case studies demonstrated how the framework can be applied to empirical transmission-pair data. In the Delta–Omicron application, the within-host-informed model supported the same qualitative conclusion as the earlier analysis of the Dutch transmission dataset [21]: Omicron had a shorter generation interval than Delta and a higher reproduction number. This finding was also consistent with other studies reporting shorter transmission intervals or altered transmission dynamics for Omicron relative to Delta [21, 27, 34–39]. The inferred generation-interval distributions could also be used directly in downstream renewal-equation analyses to estimate variant-specific reproduction numbers and the Omicron reproduction-number advantage. In the mpox application, the two within-host-informed analyses had improved predictive performance for the observed serial-interval data and supported different within-host trajectories, particularly for the decline rate and peak timing. Incorporating external mpox viral-load information from Yang et al. [30] had a clearer effect on the inferred within-host trajectory than on predictive fit to the observed serial-interval data. Thus, the empirical analyses supported the use of the framework both for estimating epidemiological delay distributions and for examining the extent to which transmission-pair data constrain their underlying biological interpretation.

The SARS-CoV-2 application likewise showed that the main advantage of the within-host-informed approach is not that it necessarily produces different epidemiological conclusions, but that the generation-interval distribution is derived from a biologically motivated transmission process rather than imposed through a prespecified parametric form. Our analysis retained the established conclusion that Omicron had a shorter generation interval than Delta [21], consistent with the shorter transmission intervals reported for Omicron in other analyses [27, 34], while providing a mechanistic explanation for the inferred delay distributions. The inferred trajectories were also broadly consistent with independent evidence for earlier Omicron transmission [21, 27, 34], although agreement with external viral-kinetic studies varied across individual within-host parameters [17, 18, 33]. Because estimates of reproduction numbers and related epidemiological quantities depend on the assumed generation-interval distribution [4, 28], improvements in the representation of transmission timing can directly affect downstream epidemiological inference.

The differences in inferred generation-interval means and dispersions across model classes highlight an important inferential advantage of the within-host-informed framework. In conventional lognormal models, the generation-interval distribution is specified directly, so features of the observed serial-interval distribution must be explained largely through changes in the mean and dispersion of that prescribed distribution. In the within-host-informed framework, generation intervals instead arise from the linked timing of infectiousness onset and subsequent transmission, allowing heterogeneity in observed transmission timing to be attributed to the latent biological processes that generate it. This can reduce dependence of the inferred generation-interval distribution on an arbitrary choice of parametric family and provide delay estimates that are more consistent with the assumed transmission process. In the SARS-CoV-2 analysis, this additional structure was associated with substantially improved predictive fit for Omicron while yielding a broader generation-interval distribution than the lognormal model. In the mpox analysis, the within-host-informed models likewise inferred substantially greater generation-interval dispersion than the lognormal comparator, despite similar fit to the observed serial-interval data. These results therefore suggest that phenomenological and mechanistic models can provide similar descriptions of observed timing data while implying materially different latent delay distributions, and that explicitly modeling the processes linking infection, infectiousness, symptoms, and transmission can provide a more principled basis for inference on those unobserved delays.

The case studies demonstrate how the framework can support applied outbreak-modeling workflows. Its primary outputs are posterior distributions for the generation interval, serial interval, and related latent delays, together with their associated uncertainty. These distributions can be used in renewal-equation estimates of reproduction numbers, variant-growth comparisons, forecasting models, and sensitivity analyses of how assumed transmission timing affects policy-relevant quantities [2, 4, 6, 28]. The framework is therefore not intended to answer intervention questions by itself, but to provide biologically structured delay distributions for downstream analyses.

Several limitations follow from the current implementation. First, the model conditions on observed transmission pairs and therefore describes the timing of realized transmission events rather than the full transmission process within a household or other risk set. It abstracts from susceptible competition, depletion, repeated exposures, non-transmission events, and variation in contact timing among potential infectees. These simplifications are not inherent to the event-time hierarchy, but they do affect how the fitted generation-interval distributions should be interpreted. The distributions estimated here are realized, observed-pair distributions: they describe the timing of transmission events represented in the household-pair data, conditional on transmission being observed. They should not be interpreted as intrinsic generation-interval distributions that would arise from the underlying infectiousness profile in an unconstrained susceptible population [1, 13].

Second, the within-host component uses a single rise-and-fall pathogen-load trajectory, whereas empirical trajectories may include multiple peaks, rebound, prolonged shedding, or richer host-level heterogeneity [15–17]. The current implementation is therefore most directly applicable to acute person-to-person infections. Environmentally mediated, vector-borne, chronic, or relapsing infections would require disease-specific components, such as environmental persistence, vector dynamics, alternative within-host trajectories, or long-term infectiousness, rather than direct use of the baseline pathogen-load/contact model.

The discrepancy between the inferred peak timing and longitudinal viral-load estimates highlights an important limitation of the current model specification. In the present framework, a single pathogen-load trajectory is assumed to drive infectiousness onset, symptom onset, and transmission, while the contact rate is held constant over time. However, the pathogen load most relevant for transmission may not coincide exactly with the trajectory measured in longitudinal clinical studies [15, 16, 32], and transmission opportunities may also change as symptoms develop and behavior is modified [40].

Third, some mechanistic parameters remain only weakly identified from transmission-pair data alone. In particular, the weak recovery of the between-host heterogeneity parameter *σ*_*z*_ indicates that serial-interval data provide only indirect information about how much variation in transmission timing arises from heterogeneity in underlying within-host trajectories. More generally, multiple mechanistic parameter combinations can imply similar and well-constrained delay distributions. Full identification of the underlying within-host parameters is therefore not required for useful estimation of epidemiological delay distributions, but mechanistic parameter estimates should be interpreted more cautiously when external biological information is unavailable.

Consequently, the appropriate level of biological interpretation depends on the inference objective. If the primary target is the epidemiological delay distribution itself, weakly informative priors may be sufficient, provided that posterior predictive fit and sensitivity to key modeling assumptions are assessed. External biological information can nevertheless sharpen inference on weakly identified components of the within-host trajectory [12]. Thus, pathogen-load, shedding, symptom-onset, incubation-period, exposure-window, or contact data should be viewed as complementary information that can strengthen biological interpretation rather than as a prerequisite for applying the framework.

The inferred delay distributions should also not be interpreted as fixed across all phases of an epidemic [2]. Some components, such as the within-host pathogen-load trajectory for a given variant and immune background, may be relatively stable over short time windows [18, 33]. However, realized generation-interval and serial-interval distributions can change with contact behavior, symptom-driven isolation, testing practices, vaccination, prior immunity, interventions, and epidemic growth rates [2, 6, 33, 40]. Where these factors vary substantially, the fitted delay distributions should therefore be interpreted as conditional on the setting and time period represented by the data.

The framework complements existing statistical and mechanistic approaches to epidemiological delay estimation. Flexible statistical delay distributions such as gamma, Weibull, and lognormal models remain useful descriptive models and provide convenient summaries of observed delay data [2, 11]. However, when fitted independently they need not correspond to a single underlying infection process or produce mutually consistent delay estimates [1, 2, 5]. Existing approaches can also represent multiple epidemiological delays jointly, but generally require parametric distributions for those delays or their component event times to be specified directly [19–21]. By contrast, the within-host-informed framework explicitly represents the biological relationships among the four key epidemiological delays, with their distributions arising from a common set of biologically motivated event hazards.

The framework is also related to contact-interval approaches in which transmission is represented through a time-to-event process [14], but here transmission is linked explicitly to the same within-host process that governs infectiousness onset and symptom onset. This distinction becomes particularly important when the inferential goal extends beyond prediction to biological interpretation. Good predictive performance for an epidemiological delay distribution does not, by itself, imply that all parameters of the latent biological process are uniquely identified. The mpox analysis illustrates this directly: the weak-prior and Yang-informed models produced similar fits to the observed serial-interval data despite supporting different combinations of peak timing and post-peak decline. External biological information can therefore be particularly valuable for constraining the mechanistic interpretation of the inferred trajectory even when it produces comparatively little change in predictive fit [12, 30].

Several natural extensions follow from the limitations of the current implementation. Because the framework is modular by construction, alternative pathogen-load curves, activation hazards, time-varying contact processes, or symptom-onset mechanisms could be substituted without changing the underlying event-time hierarchy. Jointly fitting serial-interval data with incubation-period, longitudinal pathogen-load, shedding, culture, symptom-onset, exposure-window, and contact data could improve identification of weakly constrained within-host parameters and help separate biological heterogeneity from variation in transmission opportunity [12, 30]. Allowing the contact process to vary over time through a function such as *c*(*t*) could further account for behavioral changes following symptom onset while retaining the same within-host trajectory to inform the biological event processes.

The same event-time structure could also be embedded within household risk-set or pairwise survival models so that transmission and non-transmission events, attack rates, household composition, and contact histories inform the underlying infectious-contact process [14]. Future work should therefore develop and compare joint likelihoods across these data streams, using simulation and sensitivity analyses to determine which sources of information are most valuable for specific inferential targets, including generation-interval estimation, symptom-onset timing, infectiousness timing, and biological parameter interpretation.

In conclusion, the within-host-informed event-time framework provides a principled way to construct epidemiological delay distributions from assumptions about the processes that generate infection, infectiousness, symptoms, and transmission. Rather than requiring the form of the generation-interval or related delay distributions to be selected directly from a set of convenient parametric families, the framework derives these distributions from a biologically motivated event-time model. Across the simulation studies, the resulting generation-interval distributions were recoverable across a range of sample sizes and disease time scales. In both the SARS-CoV-2 and mpox case studies, the within-host-informed models also provided competitive predictive fit to the observed timing data relative to conventional lognormal comparators. The framework therefore provides a motivated alternative to selecting epidemiological delay distributions primarily by convention, while retaining distributions that can be used directly in downstream epidemiological analyses.

## Data Availability

All data analyzed in the present study were previously published and are publicly available. SARS-CoV-2 transmission-pair and serial-interval data are available from Backer et al. (2022) and Park et al. (2023), including at:
https://pmc.ncbi.nlm.nih.gov/articles/PMC8832521/
https://github.com/parksw3/omicron-generation
https://pmc.ncbi.nlm.nih.gov/articles/PMC10235974/
Mpox transmission-pair data are publicly available from Miura et al. (2024):
https://academic.oup.com/jid/article/229/3/800/7103467
Mpox viral-load data used to construct informative priors are publicly available from Yang et al. (2024):
https://www.nature.com/articles/s41467-024-48754-8
Code used to reproduce the analyses is publicly available at:
https://github.com/epiENGAGE/Within-host-time-delay-paper

## Acknowledgments

We thank Kelly Charniga for providing feedback on an early draft of the manuscript.

## Data and code availability

The serial-interval data analyzed in this study are publicly available from the sources described in Backer et al [27] and Park et al [21]. All code and analysis used to reproduce the results presented in this study are publicly available at https://github.com/epiENGAGE/Within-host-time-delay-paper. Analyses were run in R 4.5.2 on Windows 11 using Cmd-Stan 2.38.0, `cmdstanr` 0.8.0, `posterior` 1.7.0, `dplyr` 1.2.0, `readxl` 1.4.5, `ggplot2` 4.0.1, and `loo` 2.9.0.

