## Supplementary materials for "A within-host-informed event-time framework linking epidemiological delay distributions"

### A Connections to standard delay distributions

The hazard-based formulation also clarifies how standard parametric delay distributions can arise as special cases or approximations of within-host-informed event-time models. For a generic event time $Z$ with hazard $h_Z(t)$, the distribution is determined by

$$F_Z(t) = 1 - \exp\left\{-\int_0^t h_Z(s)\,ds\right\}.$$

Different assumptions about $h_Z(t)$ therefore imply different waiting-time distributions. For example, a constant hazard gives an exponential waiting time. A hazard that changes as a power of time gives a Weibull distribution, allowing the event rate to increase or decrease with time. A hazard that changes exponentially with time gives a Gompertz distribution, corresponding to event rates that accelerate or decelerate more sharply. Additionally, a gamma distribution is naturally motivated when the observed delay is the sum of several independent and identically distributed exponential waiting times, corresponding to sequential latent stages with a common transition rate. If the sequential stages have independent exponential waiting times with different rates, the resulting delay instead follows a hypoexponential distribution.

These correspondences are useful when a fully within-host-informed model is not warranted or cannot be identified from the available data. In that setting, classical delay distributions can still be used as pragmatic observation-level models, but their choice should reflect the assumed event-time process. More generally, if classical distributions are fitted separately to different delays, they should be interpreted as approximations to particular event-time assumptions; they do not by themselves enforce consistency between the four time-delay distributions. In the present model, this activation hazard is instead driven by the pathogen-load trajectory, so the latent-period and incubation-period distributions are shaped by replication, peak timing, and clearance rather than chosen as an independent marginal distribution.

### B Observation model and censoring

To relate the derived delay distributions to real data, we introduce an observation model that accounts for censoring, partial observation, and truncation, which are common features of epidemiological delay data [1–5]. The unified formulation derived here ensures that different observation types are handled consistently within a single probabilistic framework, avoiding the need for separate models for each delay. This completes the link between the within-host-informed model and observed epidemiological data, enabling joint inference across multiple delay processes within a single coherent framework.

#### B.1 Observation model and censoring

We formulate the observation layer according to how each continuous delay variable is observed [3, 4]. Let $Z$ denote a generic latent delay variable (e.g., $IP$, $GI$, or $SI$) with density $f_Z(z \mid \Theta)$ and cumulative distribution function $F_Z(z \mid \Theta)$.

For an exactly observed continuous delay $Z_i = z_i$, the likelihood contribution is the density evaluated at the observed value,

$$L_i(\Theta) = f_Z(z_i \mid \Theta).$$

For an interval-observed delay, let $\mathcal{O}_i \subseteq \mathbb{R}$ denote the set of values of $Z_i$ consistent with the observed data. The likelihood contribution is then the probability that the latent delay lies in this observation set [2–4]:

$$L_i(\Theta) = \Pr(Z_i \in \mathcal{O}_i \mid \Theta) = \int_{\mathcal{O}_i} f_Z(z \mid \Theta)\,dz.$$

In particular, if $\mathcal{O}_i = [\ell_i, u_i]$, then

$$L_i(\Theta) = F_Z(u_i \mid \Theta) - F_Z(\ell_i \mid \Theta).$$

Together, these density and probability contributions accommodate exact observations, interval censoring, and partially observed event times.

**Incubation period.** If infection time is only known through an exposure window $I \in [E_L, E_U]$, while symptom onset is observed exactly at time $S = s$, then the incubation period satisfies

$$IP = s - I \in [s - E_U,\ s - E_L].$$

If only this implied incubation-period interval is retained, the likelihood contribution is the corresponding interval probability,

$$L_i(\Theta) = F_{IP}(s - E_L \mid \Theta) - F_{IP}(s - E_U \mid \Theta).$$

Alternatively, if infection time within the exposure window is modeled explicitly with density $g_I$, then an exactly observed symptom-onset time contributes

$$L_i(\Theta) = \int_{E_L}^{E_U} g_I(i)\, f_{IP}(s - i \mid \Theta)\, di.$$

If symptom onset is also interval observed,

$$S \in [S_L, S_U],$$

then, conditional on infection time $I = i$, the compatible incubation-period values lie in

$$IP \in [S_L - i,\ S_U - i].$$

The likelihood contribution therefore integrates the corresponding interval probability over infection-time uncertainty:

$$L_i(\Theta) = \int_{E_L}^{E_U} g_I(i)\, [F_{IP}(S_U - i \mid \Theta) - F_{IP}(S_L - i \mid \Theta)]\, di.$$

**Generation interval.** If the infection times of the infector and infectee are only known through exposure windows

$$I_1 \in [I_{1L}, I_{1U}] \qquad \text{and} \qquad I_2 \in [I_{2L}, I_{2U}],$$

then the generation interval

$$GI = I_2 - I_1$$

must lie within

$$GI \in [I_{2L} - I_{1U},\ I_{2U} - I_{1L}].$$

If only this implied generation-interval range is retained, the likelihood contribution is the corresponding interval probability,

$$L_i(\Theta) = F_{GI}(I_{2U} - I_{1L} \mid \Theta) - F_{GI}(I_{2L} - I_{1U} \mid \Theta).$$

Alternatively, if the two infection times are modeled explicitly within their respective exposure windows, with densities $g_1$ and $g_2$, then conditional on exact latent infection times $I_1 = i_1$ and $I_2 = i_2$, the generation interval is exactly

$$GI = i_2 - i_1.$$

The corresponding likelihood contribution therefore uses the generation-interval density and integrates over uncertainty in both infection times:

$$L_i(\Theta) = \int_{I_{1L}}^{I_{1U}} \int_{I_{2L}}^{I_{2U}} f_{GI}(i_2 - i_1 \mid \Theta)\, g_1(i_1)\, g_2(i_2)\, di_2\, di_1.$$

More generally, this formulation distinguishes between treating the exposure windows only through the induced interval for $GI$, which gives an interval-probability contribution, and explicitly integrating over the latent infection times, which gives a density contribution conditional on each pair of latent infection times.

**Serial interval.** If the symptom-onset times of the infector and infectee are observed exactly, then the serial interval is known exactly as

$$SI = S_2 - S_1.$$

Writing the observed serial interval as

$$s = S_2 - S_1,$$

the likelihood contribution is therefore the serial-interval density evaluated at the observed value,

$$L_i(\Theta) = f_{SI}(s \mid \Theta).$$

If symptom-onset times are instead only known within intervals

$$S_1 \in [S_{1L}, S_{1U}] \qquad \text{and} \qquad S_2 \in [S_{2L}, S_{2U}],$$

then the serial interval must lie within

$$SI \in [S_{2L} - S_{1U},\ S_{2U} - S_{1L}].$$

If only this implied serial-interval range is retained, the likelihood contribution is the corresponding interval probability,

$$L_i(\Theta) = F_{SI}(S_{2U} - S_{1L} \mid \Theta) - F_{SI}(S_{2L} - S_{1U} \mid \Theta).$$

Alternatively, if the two symptom-onset times are modeled explicitly within their respective observation windows, with densities $g_1$ and $g_2$, then conditional on exact latent onset times $S_1 = s_1$ and $S_2 = s_2$, the serial interval is exactly

$$SI = s_2 - s_1.$$

The likelihood contribution therefore uses the serial-interval density and integrates over uncertainty in both onset times:

$$L_i(\Theta) = \int_{S_{1L}}^{S_{1U}} \int_{S_{2L}}^{S_{2U}} f_{SI}(s_2 - s_1 \mid \Theta)\, g_1(s_1)\, g_2(s_2)\, ds_2\, ds_1.$$

Thus, exactly observed onset times contribute a density term for the observed serial interval, whereas interval-observed onset times can be represented either through the implied serial-interval probability or by explicitly integrating density contributions over the latent onset-time uncertainty.

#### B.2 Nested latent structure

Building on the likelihood formulation above, we now indicate how the nested latent structure enters the observation model. The serial-interval density in equation (11) of the main paper is obtained by integrating over the joint distribution of the infector's latent period, transmission delay, and incubation period, thereby preserving the dependence among these quantities induced by their shared within-host process. The resulting density $f_{SI}(s \mid \Theta)$ can be used directly for exactly observed serial intervals, while its corresponding cumulative distribution function can be used for interval-censored or truncated observations.

#### B.3 Truncation

Observation may be further distorted by truncation [1–3]. Suppose ascertainment ends at calendar time $C$, and a transmission pair is observed only if the infectee's symptom-onset time satisfies

$$S_2 \leq C.$$

If the infector's symptom-onset time $S_1 = s_1$ is observed exactly, then, because

$$SI = S_2 - S_1,$$

the ascertainment condition is equivalent to

$$SI \leq C - s_1.$$

For an exactly observed serial interval $SI = s_i$, the likelihood contribution conditional on ascertainment is therefore the right-truncated density [1–3]:

$$L_i^{\mathrm{RT}}(\Theta) = \frac{f_{SI}(s_i \mid \Theta)}{F_{SI}(C - s_{1i} \mid \Theta)}, \qquad s_i \leq C - s_{1i}.$$

For an interval-censored serial interval

$$SI \in [\ell_i, u_i],$$

the likelihood contribution is the probability of the observed interval conditional on ascertainment:

$$L_i^{\mathrm{RT}}(\Theta) = \frac{F_{SI}(\min\{u_i, C - s_{1i}\} \mid \Theta) - F_{SI}(\ell_i \mid \Theta)}{F_{SI}(C - s_{1i} \mid \Theta)},$$

provided that $\ell_i \leq C - s_{1i}$. If the observation interval is known a priori to lie entirely within the ascertainment region, so that

$$u_i \leq C - s_{1i},$$

this reduces to

$$L_i^{\mathrm{RT}}(\Theta) = \frac{F_{SI}(u_i \mid \Theta) - F_{SI}(\ell_i \mid \Theta)}{F_{SI}(C - s_{1i} \mid \Theta)}.$$

Because

$$SI = LP_1 + U_1 + IP_2 - IP_1,$$

the truncation probability under the within-host-informed model is obtained by integrating over the joint distribution of the infector-side latent event times:

$$\Pr(SI \leq C - s_1 \mid \Theta) = F_{SI}(C - s_1 \mid \Theta),$$

where

$$F_{SI}(a \mid \Theta) = \iiint f_{LP_1,U_1,IP_1}(\ell, u, i_1 \mid \Theta) F_{IP}(a - \ell - u + i_1 \mid \Theta)\, di_1\, du\, d\ell.$$

This expression preserves the dependence between the infector's latent period, transmission delay, and incubation period induced by the shared within-host process.

Truncation therefore operates at the symptom-observation level while propagating backward into inference for the latent generation-interval distribution.

If surveillance instead operates over a finite calendar-time window and pairs are observed only when

$$S_2 \in [C_0, C],$$

then, conditional on an exactly observed infector symptom-onset time $S_1 = s_1$, ascertainment is equivalent to

$$SI \in [C_0 - s_1,\ C - s_1].$$

For an exactly observed serial interval $SI = s_i$, the doubly truncated likelihood is [1, 3]

$$L_i^{\mathrm{DT}}(\Theta) = \frac{f_{SI}(s_i \mid \Theta)}{F_{SI}(C - s_{1i} \mid \Theta) - F_{SI}(C_0 - s_{1i} \mid \Theta)},$$

for

$$C_0 - s_{1i} \le s_i \le C - s_{1i}.$$

More generally, let $A_i$ denote the ascertainment event. For an exactly observed continuous delay $Z_i = z_i$, the likelihood contribution conditional on ascertainment is

$$L_i(\Theta) = f_{Z|A_i}(z_i \mid \Theta) = \frac{f_Z(z_i \mid \Theta)\,\Pr(A_i \mid Z_i = z_i, \Theta)}{\Pr(A_i \mid \Theta)}.$$

If ascertainment is deterministic given $Z_i$ and the observed value satisfies the ascertainment condition, this reduces to

$$L_i(\Theta) = \frac{f_Z(z_i \mid \Theta)}{\Pr(A_i \mid \Theta)}.$$

For an interval-observed delay $Z_i \in \mathcal{O}_i$, the corresponding conditional likelihood contribution is

$$L_i(\Theta) = \Pr(Z_i \in \mathcal{O}_i \mid A_i, \Theta) = \frac{\Pr(Z_i \in \mathcal{O}_i,\ A_i \mid \Theta)}{\Pr(A_i \mid \Theta)}.$$

If $\mathcal{O}_i$ lies entirely within the ascertainment region, this simplifies to

$$L_i(\Theta) = \frac{\displaystyle\int_{\mathcal{O}_i} f_Z(z \mid \Theta)\, dz}{\Pr(A_i \mid \Theta)}.$$

Thus, as in the untruncated observation model, exactly observed continuous delays contribute density terms, whereas interval-observed delays contribute probability terms. Truncation modifies each contribution by conditioning on the corresponding ascertainment event.

### C Case study model-fitting details

Here, we describe further details on the model-fitting procedures for the two case studies in this paper.

#### C.1 SARS-CoV-2 model-fitting details

Model fitting was performed in Stan using the `cmdstanr` interface in R. Posterior inference used Stan's dynamic Hamiltonian Monte Carlo sampler. For each fitted model, we ran four chains with 500 warmup iterations and 500 post-warmup sampling iterations per chain, giving 2,000 post-warmup posterior draws per model. We used a target average proposal acceptance probability of 0.95, allowed a maximum tree depth of 12 for each Hamiltonian Monte Carlo transition, and fixed the random-number seeds to make the analysis reproducible.

For the within-host-informed model, priors were specified on the transformed parameter scale as

$$\log \beta_I \sim \mathcal{N}(\log 0.15, 1^2), \quad \log \eta \sim \mathcal{N}(\log 0.30, 1^2),$$
$$\log r \sim \mathcal{N}(\log 0.25, 0.75^2), \quad \log d \sim \mathcal{N}(\log 0.60, 0.75^2), \quad m \sim \mathcal{N}(3.5, 1.5^2),$$
$$\log \kappa \sim \mathcal{N}(0, 1^2), \quad \log \sigma_z \sim \mathcal{N}(\log 0.35, 0.75^2), \quad \log \beta_S \sim \mathcal{N}(\log 4.0, 1^2).$$

These priors were chosen to be weakly informative on the time scales plausible for acute respiratory-virus transmission, while allowing substantial uncertainty in pathogen-load growth, clearance, peak timing, infectiousness activation, symptom-onset activation, and between-host heterogeneity.

For the lognormal comparison model, priors were placed on the lognormal generation-interval mean and standard deviation parameters using the specification in [5]. Model comparison was performed using pointwise log-likelihood values generated in Stan, from which WAIC and PSIS-LOO were computed using the `loo` package. Posterior predictive serial-interval distributions were generated by evaluating the fitted serial-interval probabilities over the observed support and drawing replicated datasets with the same sample size as the corresponding strain-specific household transmission dataset.

#### C.2 Mpox model fitting details

The mpox analysis used publicly available transmission-pair data from [6]. We restricted the analysis to high-confidence transmission pairs with transmission-pair confidence score equal to 9, high confidence in the transmission link, and high confidence in symptom-onset dates. This restriction yielded 34 transmission pairs. Serial intervals were calculated as the difference between the symptom-onset date of the infectee and the symptom-onset date of the infector. Repeated serial-interval values were aggregated into weighted counts for model fitting.

##### C.2.1 Lognormal generation-interval comparator

The phenomenological comparator specified the generation interval directly as a lognormal distribution. The serial-interval likelihood was obtained by combining this generation-interval distribution with a fixed lognormal incubation-period distribution. The incubation-period distribution was assigned mean 8.5 days and standard deviation 4.5 days, converted internally to lognormal parameters. The epidemic-growth correction was set to $r_{\text{growth}} = 0$ for the standalone mpox comparator, and the correlation between the log incubation period of the infector and the log generation interval was set to $\rho = 0$.

Priors for the lognormal generation-interval model were specified on the lognormal parameters. The prior center was defined by a generation-interval mean of 12 days and standard deviation of 7 days, converted to lognormal parameters. The prior standard deviations were

$$\sigma_{\mu_{\text{gen}}} = 1.0, \qquad \sigma_{\sigma_{\text{gen}}} = 0.75,$$

where $\mu_{\text{gen}}$ and $\sigma_{\text{gen}}$ denote the lognormal location and scale parameters for the generation interval.

The serial-interval normalization grid used spacing 0.25 days over $[-30, 60]$ days. The incubation-period quadrature grid used spacing 0.25 days over $[0.05, 45]$ days.

##### C.2.2 Weak-prior within-host-informed mpox model

The weak-prior within-host-informed model used the same observed-pair event-time structure as the main framework, fitted to the 34 high-confidence mpox transmission pairs. The model used a neutral epidemic-growth correction with $r_{\text{growth}} = 0$. Because mpox serial intervals in this dataset were longer than in the SARS-CoV-2 application, wider numerical grids were used. The serial-interval grid used spacing 0.25 days over $[-15, 40]$ days. The latent-period and post-infectiousness transmission-delay grids used spacing 0.25 days over $[0, 45]$ days. The generation-interval grid extended to 90 days, and the incubation-period grid used spacing 0.05 days over $[0.05, 60]$ days. Between-host heterogeneity in the within-host trajectory was integrated using 11 quadrature points over $\pm 3.5$ standard deviations.

For the weak-prior within-host-informed model, priors were specified on transformed parameters as

$$\log \beta_I \sim \mathcal{N}(\log 0.08, 1.5^2), \qquad \log \beta_S \sim \mathcal{N}(\log 0.08, 1.5^2),$$
$$\log r \sim \mathcal{N}(\log 0.15, 1.0^2), \qquad \log d \sim \mathcal{N}(\log 0.20, 1.0^2), \qquad m \sim \mathcal{N}(8.0, 5.0^2),$$
$$\log \kappa \sim \mathcal{N}(0, 1.25^2), \qquad \log \sigma_z \sim \mathcal{N}(\log 0.50, 1.0^2).$$

These priors were intended to place the model on a plausible mpox time scale while remaining broad enough to avoid imposing viral-load-derived information in the weak-prior analysis.

##### C.2.3 Yang-informed viral-load prior construction

To construct viral-load-informed priors, we fitted the same within-host trajectory form used in the event-time model to mpox viral-load observations from [7]. The fitted trajectory had the form

$$V(t) = \exp(rt) \left\{1 + \frac{r}{d} \exp[\kappa(t - m)]\right\}^{-(r+d)/\kappa},$$

where $t$ denotes time since symptom onset in the viral-load dataset. The viral-load model estimated posterior distributions for $r$, $d$, $m$, and $\kappa$, with weak regularizing priors

$$\log r \sim \mathcal{N}(\log 0.35, 1.0^2), \qquad \log d \sim \mathcal{N}(\log 0.20, 1.0^2),$$
$$m \sim \mathcal{N}(6.0, 4.0^2), \qquad \log \kappa \sim \mathcal{N}(0, 1.0^2).$$

Values reported at the lower detection limit, $y = 3$ on the $\log_{10}$ scale, were treated as left-censored by default. The viral-load model included site-specific intercepts and an HIV-status effect. Because the public source data did not provide participant identifiers for the Figure 2 viral-load observations, the viral-load analysis estimated population-level trajectory shape rather than the individual-level heterogeneity parameter $\sigma_z$ used in the transmission-pair model.

The Yang-informed prior was constructed from posterior draws of

$$(\log r, \log d, m, \log \kappa).$$

Because the viral-load data were indexed by time since symptom onset, whereas the event-time transmission model uses time since infection, the posterior draws for $m$ were shifted to the infection-time scale by adding an incubation-period offset. The incubation-period offset had mean 8 days and standard deviation 3 days. The shifted posterior draws were then converted into a multivariate normal approximation with mean $\mu_{\text{Yang}}$ and covariance matrix $\Sigma_{\text{Yang}}$, preserving posterior covariance among the trajectory parameters.

##### C.2.4 Yang-informed robust-prior transmission-pair model

The Yang-informed within-host-informed transmission-pair model used an informative multivariate normal prior for

$$\theta = (\log r, \log d, m, \log \kappa).$$

The prior was

$$p(\theta) = \mathcal{N}(\theta \mid \mu_{\text{Yang}}, \Sigma_{\text{Yang}}).$$

Before fitting the transmission-pair model, the Yang-derived covariance matrix was inflated by a factor of 2 on the standard-deviation scale. This robust-prior formulation allowed the model to borrow information from external viral-load data while retaining support for broader within-host trajectories if the viral-load and transmission-pair data favored incompatible timing structures. The event-hazard scale parameters and $\sigma_z$ were left under weak priors, because the Yang viral-load data do not directly identify infectiousness activation, symptom-onset activation, transmission-pair timing, or between-host heterogeneity in the transmission model.

##### C.2.5 Computation and model comparison

All mpox models were fitted in Stan using the `cmdstanr` interface in R. For the lognormal and weak-prior within-host-informed transmission-pair models, we ran four chains with 500 warmup iterations and 500 post-warmup sampling iterations per chain, giving 2,000 post-warmup posterior draws per model. The target average proposal acceptance probability was set to 0.95 and the maximum tree depth was set to 12.

The Yang viral-load model was fitted using four chains with 1,000 warmup iterations and 1,000 post-warmup sampling iterations per chain. The target average proposal acceptance probability was set to 0.95 and the maximum tree depth was set to 12.

For each fitted transmission-pair model, pointwise log-likelihood values were generated in Stan. Predictive performance was assessed using WAIC and PSIS-LOO computed from these pointwise log-likelihood values using the `loo` package. Posterior summaries were computed for the generation-interval mean and standard deviation, the within-host trajectory parameters, and the event-hazard scale parameters. For the viral-load-informed analysis, posterior draws from the Yang-informed prior were compared with the weak-prior transmission-pair posterior for $(\log r, \log d, m, \log \kappa)$, to assess whether the within-host trajectories favored by serial-interval data alone were compatible with those implied by external viral-load measurements.

### D Simulation study details

This appendix provides additional details for the four simulation analyses described in Section 2.4. All simulations used the observed-pair version of the within-host-informed serial-interval model. In this formulation, the post-infectiousness transmission delay $U_1$ is interpreted as the timing of a selected observed transmission event, conditional on a transmission pair being observed. Therefore, conditional on the latent period, the density of $U_1$ is proportional to the pathogen-load-driven transmission intensity and normalized over the transmission-delay grid. The absolute transmission-intensity scale cancels from this conditional timing density and is not identifiable from serial-interval pair timing alone.

All simulations used the pathogen-load trajectory

$$V(t) = \exp(rt)\left\{1 + \frac{r}{d}\exp[\kappa(t-m)]\right\}^{-(r+d)/\kappa},$$

where $r$ is the early growth rate, $d$ is the late decay rate, $m$ is the peak time, and $\kappa$ controls the sharpness of the transition between growth and decay. This parameterization makes $m$ the exact peak time. Infectiousness onset and symptom onset were generated using proportional hazards of the form $\beta_I V(t)$ and $\beta_S V(t)$, respectively, with multiplicative between-host heterogeneity represented through $\sigma_z$.

#### D.1 Simulation 1: finite-sample recovery

The first simulation assessed whether the model could recover broad generation-interval summaries from small transmission-pair datasets. Data were generated from a single fixed parameter set:

$$\log \beta_I = \log(0.16), \quad \log \beta_S = \log(0.16), \quad \log r = \log(0.27), \quad m = 3.5,$$
$$\log d = \log(0.62), \quad \log \kappa = \log(1.05), \quad \log \sigma_z = \log(0.35).$$

For each sample size $n \in \{30, 50, 100\}$, we generated 100 independent replicate datasets by multinomial sampling from the true model-implied serial-interval probability mass function. Each simulated dataset was refitted using the same observed-pair within-host-informed model. The prior distributions were:

$$\log \beta_I \sim N\{\log(0.15), 1.0^2\}, \quad \log \beta_S \sim N\{\log(0.15), 1.0^2\},$$

$$\log r \sim N\{\log(0.25), 0.75^2\}, \quad \log d \sim N\{\log(0.60), 0.75^2\},$$

$$m \sim N(3.5, 1.5^2), \quad \log \kappa \sim N(0, 1.0^2), \quad \log \sigma_z \sim N\{\log(0.35), 0.75^2\}.$$

These priors were deliberately centered close to the data-generating parameter values. In particular, the prior centers for $m$ and $\sigma_z$ were exactly equal to their generating values, while the prior centers for $\beta_I$, $\beta_S$, $r$, $d$, and $\kappa$ were also close to the corresponding generating values. Accordingly, this simulation should be interpreted as a well-specified recovery experiment rather than as an assessment of how much information about the generation-interval distribution is supplied by the serial-interval data alone. This distinction is particularly important at $n = 30$, where the favorably centered priors may contribute materially to posterior recovery. Sensitivity to prior accuracy and strength is examined separately in Simulation 3. The parameter bounds used in Stan were retained during fitting.

Model fitting used four Markov chains, 250 warmup iterations and 250 sampling iterations per chain, `adapt_delta` $= 0.99$, and maximum tree depth 15. The numerical grids used spacing 0.5 days for serial intervals, latent periods, transmission delays, and generation intervals, and 0.10 days for incubation periods. The serial-interval support was $[-10, 20]$ days, the latent-period and transmission-delay supports were $[0, 20]$ days, the generation-interval support was $[0, 40]$ days, and the incubation-period support was $[0.05, 30]$ days. Between-host heterogeneity was integrated using seven quadrature points spanning $\pm 3.5$ standard deviations.

For each fit, we recorded posterior medians and 90% posterior intervals for the generation-interval mean and standard deviation. Performance was summarized using bias, absolute error, root mean squared error, posterior interval width, empirical coverage of the known data-generating value, convergence diagnostics, and effective sample sizes. We also summarized recovery of the underlying event-time parameters, but interpreted this as secondary because the primary aim was recovery of epidemiological delay summaries rather than full mechanistic parameter identification.

#### D.2 Simulation 2: recovery across generation-interval regimes

The second simulation assessed whether the within-host-informed observed-pair model could recover generation-interval summaries across infections with substantially different time scales. The aim was to test whether the framework was specific to short-generation-interval infections, such as SARS-CoV-2, or whether it could also recover delay distributions in intermediate- and long-generation-interval regimes.

We considered three data-generating regimes with target mean generation intervals of approximately 3, 10, and 20 days. For each regime, we specified an initial within-host-informed parameter set and then numerically tuned the infectiousness-onset hazard scale so that the model-implied mean generation interval was close to the target value. Because the observed-pair transmission-delay density is conditioned on an observed transmission event, the absolute transmission-intensity scale cancels from the timing likelihood; therefore, calibration was performed by shifting the infectiousness-onset hazard scale.

The initial regime parameter sets were

| Regime | $\log\beta_I$ | $\log\beta_S$ | $\log r$ | $\log d$ | $m$ | $\log\kappa$ | $\log\sigma_z$ |
|---|---|---|---|---|---|---|---|
| Mean 3 | log(0.16) | log(0.16) | log(0.27) | log(0.62) | 3.5 | log(1.05) | log(0.35) |
| Mean 10 | log(0.06) | log(0.10) | log(0.18) | log(0.25) | 7.5 | log(0.70) | log(0.35) |
| Mean 20 | log(0.025) | log(0.08) | log(0.12) | log(0.13) | 11.0 | log(0.45) | log(0.35) |

where $\beta_I$ is the infectiousness-onset hazard scale, $\beta_S$ is the symptom-onset hazard scale, $r$ is the early pathogen-load growth rate, $d$ is the post-peak decline rate, $m$ is the peak time, $\kappa$ controls the sharpness of the transition from growth to decline, and $\sigma_z$ controls between-host heterogeneity.

For each regime, the calibrated parameter set was used to compute the true model-implied serial-interval distribution and generation-interval distribution. We then simulated $n = 200$ serial intervals from the corresponding serial-interval distribution. This was repeated for 5 simulation replicates per regime. Because this simulation was intended as a regime-recovery check rather than a high-precision Monte Carlo assessment, we used five replicates per regime. The epidemic-growth correction was set to $r_{\text{growth}} = 0$, so the simulation assessed recovery of the intrinsic observed-pair timing distribution without epidemic-growth distortion.

All simulated datasets were refitted using the same within-host-informed observed-pair model. To avoid favoring a SARS-CoV-2-like time scale, we used broad common priors for all three regimes:

$$\log\beta_I \sim \mathcal{N}(\log 0.10, 2.0^2), \qquad \log\beta_S \sim \mathcal{N}(\log 0.12, 2.0^2),$$

$$\log r \sim \mathcal{N}(\log 0.20, 1.25^2), \qquad \log d \sim \mathcal{N}(\log 0.30, 1.25^2), \qquad m \sim \mathcal{N}(8.0, 6.0^2),$$

$$\log\kappa \sim \mathcal{N}(\log 0.75, 1.25^2), \qquad \log\sigma_z \sim \mathcal{N}(\log 0.35, 1.0^2).$$

These priors were chosen to place the short, intermediate, and long-generation-interval regimes within prior support rather than imposing the more concentrated SARS-CoV-2 prior structure used in the empirical analysis.

The numerical grids were widened relative to the small-sample simulation to accommodate the long-generation-interval regime. The serial-interval grid used spacing 1.0 day over $[-30, 80]$ days. The latent-period and post-infectiousness transmission-delay grids used spacing 1.0 day over $[0, 90]$ days. The generation-interval grid extended to 180 days. The incubation-period grid used spacing 0.25 days over $[0.05, 70]$ days. Between-host heterogeneity in the within-host trajectory was integrated using 5 quadrature points over $\pm 3.5$ standard deviations.

The Stan model used wider parameter bounds than the small-sample simulation to allow the long-generation-interval regimes to be fitted. The bounds were

$$\log\beta_I \in [-8, 8], \qquad \log\beta_S \in [-8, 8],$$

$$\log r \in [-5, 2], \qquad \log d \in [-5, 2], \qquad m \in [0.25, 12],$$

$$\log\kappa \in [-5, 3], \qquad \log\sigma_z \in [-6, 1.5].$$

All models were fitted in Stan using `cmdstanr`. For each fitted dataset, we ran four chains with 500 warmup iterations and 500 post-warmup sampling iterations per chain, giving 2,000 post-warmup posterior draws per fitted model. The target average proposal acceptance probability was set to 0.99, and the maximum tree depth was set to 15.

For each fitted model, we summarized recovery of the generation-interval mean and standard deviation using posterior medians and central 90% posterior intervals. Performance was evaluated using bias, absolute error, root mean squared error, posterior interval width, posterior interval coverage, convergence diagnostics, and fit failures. The main text reports posterior median estimates and absolute percentage errors for the generation-interval mean and standard deviation across the three regimes.

##### D.3 Simulation 3: prior sensitivity and within-host parameter learnability

The third simulation assessed how inference from serial-interval data alone depends on the accuracy and strength of prior information about the within-host trajectory. The aim was to distinguish recovery of observed-scale epidemiological delay summaries from identification of the latent within-host-informed parameters.

Data were generated from the observed-pair within-host-informed serial-interval model using the fixed parameter set

$$\log\beta_I = \log(0.16), \qquad \log\beta_S = \log(0.16),$$

$$\log r = \log(0.27), \qquad \log d = \log(0.62), \qquad m = 3.5,$$

$$\log\kappa = \log(1.05), \qquad \log\sigma_z = \log(0.35).$$

The epidemic-growth correction was set to $r_{\text{growth}} = 0$. For each simulation replicate, we generated a training dataset of $n = 200$ serial intervals and an independent held-out test dataset of $n_{\text{test}} = 5000$ serial intervals from the true model-implied serial-interval distribution. We used 50 simulation replicates. Within each replicate, the same simulated training and test datasets were used for all prior scenarios.

We considered prior scenarios that varied both prior accuracy and prior strength. Prior perturbations were applied to the within-host trajectory parameters

$$(\log r, \log d, m, \log\kappa, \log\sigma_z),$$

which represent the parameters most plausibly informed by external pathogen-load or shedding data. The event-hazard scale parameters $\log\beta_I$ and $\log\beta_S$ were kept under the manuscript-default broad priors in all scenarios, because pathogen-load data inform the trajectory shape but do not directly identify the conversion from pathogen load to infectiousness or symptom onset.

The default priors were

$$\log\beta_I \sim \mathcal{N}(\log 0.15, 1.0^2), \qquad \log\beta_S \sim \mathcal{N}(\log 0.15, 1.0^2),$$

$$\log r \sim \mathcal{N}(\log 0.25, 0.75^2), \qquad \log d \sim \mathcal{N}(\log 0.60, 0.75^2), \qquad m \sim \mathcal{N}(3.5, 1.5^2),$$

$$\log\kappa \sim \mathcal{N}(0, 1.0^2), \qquad \log\sigma_z \sim \mathcal{N}(\log 0.35, 0.75^2).$$

For the prior-sensitivity grid, prior accuracy was classified as accurate, mildly wrong, or badly wrong. Accurate priors were centered on the true data-generating values. Mildly wrong priors used offsets

$$\Delta\log r = \log(1.25), \qquad \Delta\log d = \log(0.85), \qquad \Delta m = 0.45,$$

$$\Delta\log\kappa = \log(1.25), \qquad \Delta\log\sigma_z = \log(1.20),$$

relative to the true values. Badly wrong priors used offsets

$$\Delta\log r = \log(1.60), \qquad \Delta\log d = \log(0.65), \qquad \Delta m = 0.90,$$

$$\Delta \log \kappa = \log(1.80), \qquad \Delta \log \sigma_z = \log(1.60).$$

Prior strength was classified as weak, moderate, or strong. For log-scale trajectory parameters, the prior standard deviations were 0.75, 0.35, and 0.15, respectively. For $m$, the corresponding prior standard deviations were 1.50, 0.60, and 0.25 days. The full simulation therefore crossed three prior-accuracy settings with three prior-strength settings, with an additional manuscript-default prior scenario.

The numerical grids were chosen to match the time scale of the data-generating distribution while keeping repeated simulation fitting computationally feasible. The serial-interval grid used spacing 0.5 days over $[-10, 20]$ days. The latent-period and post-infectiousness transmission-delay grids used spacing 0.5 days over $[0, 20]$ days. The generation-interval grid extended to 40 days. The incubation-period grid used spacing 0.10 days over $[0.05, 30]$ days. Between-host heterogeneity in the within-host trajectory was integrated using 7 quadrature points over $\pm 3.5$ standard deviations.

All models were fitted in Stan using `cmdstanr`. For each replicate and prior scenario, we ran four chains with 300 warmup iterations and 300 post-warmup sampling iterations per chain, giving 1,200 post-warmup posterior draws per fitted model. The target average proposal acceptance probability was set to 0.99, and the maximum tree depth was set to 15.

For each fitted model, we summarized recovery of the generation-interval mean and standard deviation using posterior medians and central 90% posterior intervals. We computed absolute errors for the generation-interval mean and standard deviation relative to the true model-implied values. We also evaluated observed-scale recovery by comparing the posterior mean serial-interval distribution with the true serial-interval distribution using Kullback–Leibler distance, total-variation distance, and root mean squared error across serial-interval grid cells. Held-out predictive performance was assessed using the independent test dataset, reporting held-out log predictive density per pair. WAIC and PSIS-LOO were also computed from pointwise log-likelihood values where applicable.

To assess parameter learnability, we compared the posterior median of each within-host-informed parameter with both the true value and the prior mean. For each parameter and scenario, we recorded whether the posterior median moved closer to the true value than the prior mean. This quantity was summarized across simulation replicates for the trajectory parameters

$$(\log r, \log d, m, \log \kappa, \log \sigma_z),$$

and was used to assess which mechanistic parameters were updated by serial-interval data and which remained prior-dominated.

#### D.4 Simulation 4: joint recovery of epidemiological delays

The fourth simulation assessed whether the within-host-informed observed-pair model could simultaneously recover multiple epidemiological delay distributions from serial-interval data alone. The aim was to evaluate whether the latent period, incubation period, generation interval, and serial interval can be inferred jointly as linked outcomes of the same underlying event-time process, rather than being estimated as separate marginal distributions.

Data were generated using the same SARS-CoV-2-like parameter set as in Simulation 1:

$$\log \beta_I = \log(0.16), \qquad \log \beta_S = \log(0.16),$$

$$\log r = \log(0.27), \qquad \log d = \log(0.62), \qquad m = 3.5,$$

$$\log \kappa = \log(1.05), \qquad \log \sigma_z = \log(0.35).$$

Under this data-generating model, the true model-implied latent-period distribution had mean 2.61 days and standard deviation 2.05 days, while the incubation-period distribution had mean 2.74 days and standard deviation 2.00 days. The corresponding generation-interval distribution had mean 5.06 days and standard deviation 2.43 days, and the serial-interval distribution had mean 5.05 days and standard deviation 3.68 days. These quantities were calculated directly from the known data-generating model using the same numerical integration scheme used for model fitting.

We generated 100 independent replicate datasets, each containing $n = 150$ serial intervals sampled from the true model-implied serial-interval probability mass function. Only the simulated serial intervals were supplied to the fitted model; the latent periods, incubation periods, and generation intervals used as recovery targets were not included as observed data. Each replicate was refitted using the same observed-pair within-host-informed model as in Simulation 1.

The prior distributions were also the same as those used in Simulation 1:

$$\log \beta_I \sim \mathcal{N}\{\log(0.15), 1.0^2\}, \qquad \log \beta_S \sim \mathcal{N}\{\log(0.15), 1.0^2\},$$
$$\log r \sim \mathcal{N}\{\log(0.25), 0.75^2\}, \qquad \log d \sim \mathcal{N}\{\log(0.60), 0.75^2\},$$
$$m \sim \mathcal{N}(3.5, 1.5^2), \qquad \log \kappa \sim \mathcal{N}(0, 1.0^2), \qquad \log \sigma_z \sim \mathcal{N}\{\log(0.35), 0.75^2\}.$$

The numerical grids matched those used in Simulation 1. Serial intervals were represented on a 0.5-day grid over $[-10, 20]$ days. The latent-period and post-infectiousness transmission-delay grids used spacing 0.5 days over $[0, 20]$ days, while the generation-interval grid extended from 0 to 40 days with the same spacing. The incubation-period grid used spacing 0.10 days over $[0.05, 30]$ days. Between-host heterogeneity was integrated using seven quadrature points spanning $\pm 3.5$ standard deviations.

All models were fitted in Stan using `cmdstanr`. For each replicate, we ran four chains with 250 warmup iterations and 250 post-warmup sampling iterations per chain, giving 1,000 post-warmup posterior draws per fitted model. The target average proposal acceptance probability was set to 0.99, and the maximum tree depth was set to 15. To reduce computational cost without changing the statistical model, posterior reconstruction of the epidemiological delay distributions was performed after sampling using standalone generated quantities evaluated at the posterior draws.

For each posterior draw, we derived the model-implied latent-period, incubation-period, generation-interval, and serial-interval distributions. For each of the four delays, we summarized the posterior distribution of its mean and standard deviation. Recovery of these eight scalar targets was evaluated using bias and absolute error of the posterior median, root mean squared error across simulation replicates, continuous ranked probability score (CRPS) for the full posterior distribution, width of the central 90% posterior interval, and empirical coverage of the known data-generating value.

We additionally evaluated recovery of the complete delay distributions rather than only their means and standard deviations. For each replicate, the fitted latent-period, incubation-period, generation-interval, and serial-interval probability distributions were compared with their corresponding known data-generating distributions using total-variation distance, root mean squared error across probability-mass grid cells, and Kullback–Leibler divergence from the true to the fitted distribution. This allowed the simulation to assess whether the model recovered not only scalar summaries but also the shapes of all four epidemiological delay distributions jointly.

### E Extra SARS-CoV-2 analyses

#### E.1 Serial interval distributions

The within-host-informed models showed the clearest visual improvement for Omicron, while for Delta the censored within-host-informed and lognormal models fitted the peak similarly well and the within-host-informed models better captured the left tail.

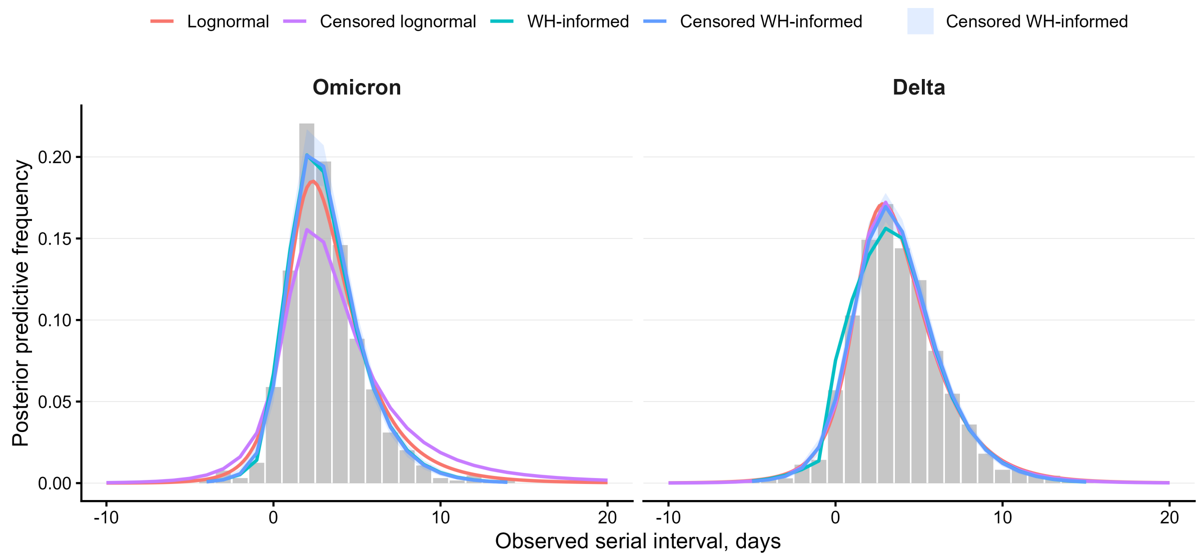


**Figure S1: Observed and fitted serial-interval distributions for Omicron ($n = 644$) and Delta ($n =$ 1,333) household transmission pairs.** Posterior predictive serial-interval distributions were generated by simulating transmission pairs from the fitted models using the observed sample sizes for each variant. Gray bars show the empirical observed serial-interval frequencies. Colored curves show posterior median fitted frequencies for the lognormal, censored lognormal, within-host-informed, and censored within-host-informed models, with shaded regions denoting the corresponding 95% posterior predictive intervals.

For both Delta, all four models reproduced the main features of the observed serial-interval distributions, including the modal interval and the decline in frequency at longer serial intervals. For Omicron, both within-host-informed models captured the observed peak more accurately than the lognormal models while also providing a better fit to both tails of the distribution. For Delta, the lognormal model and the censored within-host-informed model reproduced the observed peak similarly well; these two models provided similar fits to the right tail. These visual differences are consistent with the improved predictive performance of the within-host-informed models observed in the WAIC and LOOIC analyses.

##### E.1.1 Effect of generation-interval shape on reproduction-number estimation

We compared reproduction-number estimates obtained using the within-host-informed and lognormal realized generation-interval distributions (Figure S2). Because the two models inferred broadly similar mean generation intervals, large differences in the temporal pattern of $R_v(t)$ are not expected. Instead, any differences primarily reflect how the two generation-interval distributions allocate probability mass across transmission lags, thereby altering the renewal denominator.

The within-host-informed estimates suggest that the Omicron reproduction number declined from approximately 1.5–1.6 in early December to around 1.2 by late January. In contrast, the Delta reproduction number remained below the epidemic threshold throughout, decreasing from approximately 0.8 in early December to around 0.65–0.7 in early January before increasing slightly toward the end of the period. Consequently, the inferred Omicron reproduction-number advantage was initially close to two-fold and declined toward approximately 1.4 by late January. This pattern is consistent with the changing growth-rate advantage of Omicron during variant replacement.

Replacing the lognormal generation-interval distribution with the within-host-informed distribution had only a modest effect on the inferred reproduction numbers and did not alter their qualitative temporal behavior. The within-host-informed estimates are generally slightly higher than the corresponding lognormal estimates, most visibly for Omicron and for the Omicron-to-Delta reproduction-number advantage. These differences reflect changes in the shape of the generation-interval distribution rather than its mean, which subtly alter how past infections contribute to the renewal denominator. Although the effect was modest in this application, it may become more important when transmission timing changes over time, such as during variant replacement or shifts in population behavior.

Importantly, both approaches led to the same substantive conclusion as [5]: allowing Delta and Omicron to have different generation-interval distributions is important for estimating differences in their reproduction numbers.

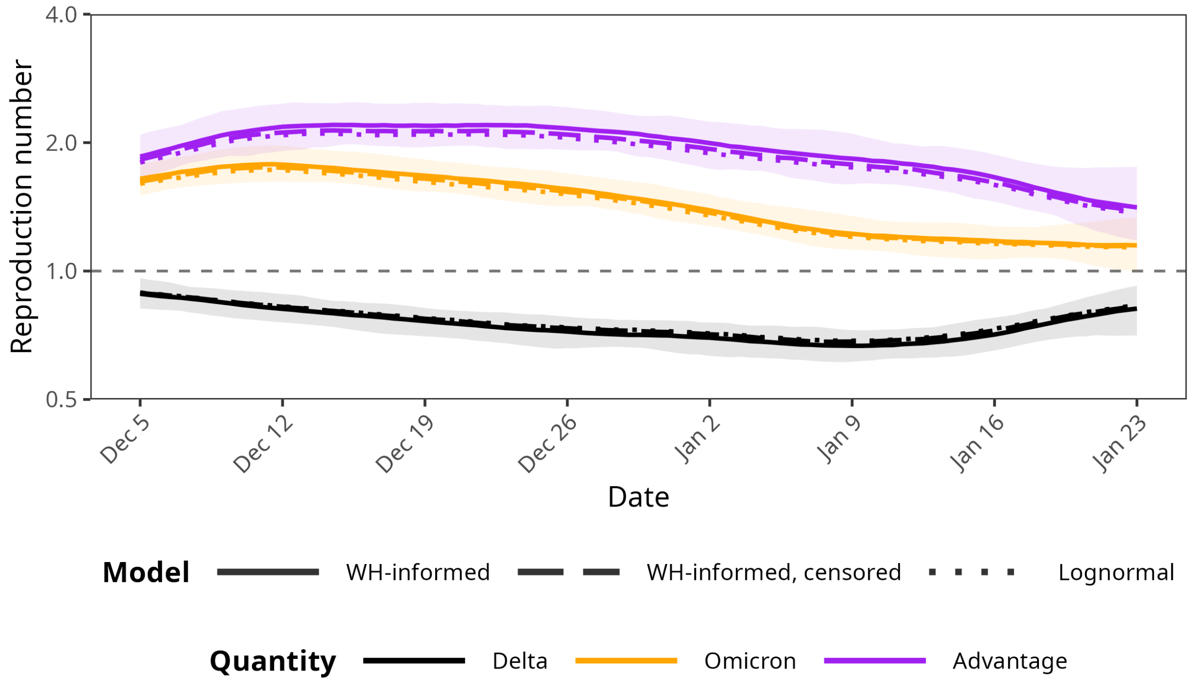


**Figure S2: Estimated reproduction numbers obtained using the within-host-informed and lognormal realized generation-interval distributions.** Solid lines and shaded regions show estimates obtained from the within-host-informed model; dotted lines show estimates from the lognormal model. Black denotes Delta, orange denotes Omicron, and purple denotes the ratio of the Omicron and Delta instantaneous reproduction numbers. The horizontal dashed line indicates the epidemic threshold ($R = 1$).
